# *Streptococcus* species abundance in the gut is linked to subclinical coronary atherosclerosis in 8973 participants from the SCAPIS cohort

**DOI:** 10.1101/2022.05.25.22275561

**Authors:** Sergi Sayols-Baixeras, Koen F. Dekkers, Gabriel Baldanzi, Daniel Jönsson, Ulf Hammar, Yi-Ting Lin, Shafqat Ahmad, Diem Nguyen, Georgios Varotsis, Sara Pita, Nynne Nielsen, Aron C. Eklund, Jacob B. Holm, H. Bjørn Nielsen, Ulrika Ericson, Louise Brunkwall, Filip Ottosson, Anna Larsson, Dan Ericson, Björn Klinge, Peter M. Nilsson, Andrei Malinovschi, Lars Lind, Göran Bergström, Johan Sundström, Johan Ärnlöv, Gunnar Engström, J. Gustav Smith, Marju Orho-Melander, Tove Fall

## Abstract

**BACKGROUND:** Gut microbiota have been implicated in atherosclerotic disease, but their relation with subclinical coronary atherosclerosis is unclear. This study aimed to identify associations between the gut microbiome and computed tomography-based measures of coronary atherosclerosis, and to explore relevant clinical correlates.

**METHODS:** We conducted a cross-sectional study of 8973 participants aged 50 to 65 without overt atherosclerotic disease from the population-based Swedish Cardiopulmonary BioImage Study (SCAPIS). Coronary atherosclerosis was measured using coronary artery calcium score (CACS) and coronary computed tomography angiography (CCTA). Gut microbiota species abundance and functional potential were assessed with shotgun metagenomics sequencing of stool samples, and their association with coronary atherosclerosis was evaluated with multivariable regression models adjusted for cardiovascular risk factors. Associated species were evaluated for association with inflammatory markers, metabolites, and corresponding species in saliva.

**RESULTS:** The mean age of the study sample was 57.4 years, and 53.7% were female. Coronary artery calcification was detected in 40.3% of participants, and 5.4% had at least one stenosis with more than 50% occlusion. Sixty-four species were associated with CACS independent of cardiovascular risk factors, with the strongest associations observed for *Streptococcus anginosus* and *S. oralis* subsp*. oralis* (*P*<1×10^-5^). Associations were largely similar across CCTA-based measurements. Out of the 64 species, 19 species, including streptococci and other species commonly found in the oral cavity, were associated with high-sensitivity C-reactive protein plasma concentrations and 16 with neutrophil counts. Oral species in the gut were negatively associated with plasma indole propionate and positively associated with plasma secondary bile acids and imidazole propionate. Five species correlated with the same species in saliva and were associated with worse dental health in the Malmö Offspring Dental Study. Microbial functional potential of dissimilatory nitrate reduction, anaerobic fatty acid beta-oxidation and amino acid degradation was associated with CACS.

**CONCLUSIONS:** This study provides evidence of an association of a gut microbiota composition characterized by increased abundance of *Streptococcus* spp. and other species commonly found in the oral cavity with coronary atherosclerosis and systemic inflammation. Further longitudinal and experimental studies are warranted to explore the potential implication of a bacterial component in atherogenesis.

**CLINICAL PERSPECTIVE:** *WHAT IS NEW?:*

- Shotgun metagenomics identified associations between gut species and subclinical atherosclerosis assessed with computed tomography-derived coronary artery calcium score (CACS) in 8973 participants, with an overrepresentation of the *Streptococcus* and *Oscillobacter* genera.
- The relative abundance of CACS-associated oral species detected in fecal samples was negatively associated with indole propionate, while positively associated with secondary bile acids and imidazole propionate.
- Gut *Streptococcus* spp. were positively associated with circulating biomarkers of systemic inflammation and infection response, and with the same species located in the mouth, which were in turn associated with oral pathologies.

*WHAT ARE THE CLINICAL IMPLICATIONS?:*

- We describe the link between gut microbiota composition, especially species commonly found in the mouth, with subclinical coronary atherosclerosis and biomarkers of inflammation in the largest cardiovascular and metagenomics study to date.
- The effects of gut and oral *Streptococcus* spp. on risk for coronary artery disease merit further longitudinal and experimental studies.

---

Atherosclerotic cardiovascular disease (CVD) is a major cause of death and disability. ^1^ The microbial community of the gastrointestinal tract, also known as the gut microbiota, is hypothesized to affect the progression of atherosclerosis through three potential mechanisms. Firstly, microbial metabolites can interfere with the host metabolism, including lipid metabolism. ^2^ The composition of the gut microbiota has been linked to metabolic disorders such as obesity, insulin resistance and type 2 diabetes, although the causal relation and direction are unclear. ^3^ Secondly, translocation of live bacteria or bacterial structural components (e.g., endotoxins) into the bloodstream can contribute to low-grade systemic inflammation, thereby exacerbating the atherosclerosis process. ^2^ Thirdly, the discovery of bacteria within atherosclerotic plaques has led to the proposal that bacteria might directly infect plaques and accelerate atherosclerosis progression. ^4^ Case-control studies of symptomatic coronary atherosclerotic disease have identified differences in the abundance of more than 500 gut species. ^5–7^ However, dissimilarities in medical treatment and lifestyle factors in cases and controls make these studies prone to bias. Thus, studies of individuals without overt coronary disease in large population-based samples incorporating biomarkers of metabolism and inflammation are warranted.

Recent evidence supports that oral species are commonly transmitted to the gut, ^8^ indicating that the gut and oral microbiota are connected rather than being two separate microbial communities. Causative species of dental caries, plaque-dwelling bacteria and endocarditis-associated species including species from the viridans streptococci group (VGS) are reported to be transmitted at a high rate. Additionally, an overlap is reported between oral bacteria such as *Veillonella* spp. and *Streptococcus* spp. found in oral cavity, fecal samples and carotid atherosclerotic plaque. ^9^ Given the association between dental health and endothelial dysfunction and atherosclerotic disease, ^10–12^ the gut may serve as a niche and/or entry route for oral pathogenic bacteria passing to the blood. However, there is a lack of studies assessing the association of atherosclerosis-related gut species with the oral microbiome.

To avoid the limitations of previous studies and gain insight into the relation of the gut microbiota and atherosclerosis, we sought to identify associations between the gut microbiome and computed tomography-based measures of subclinical coronary atherosclerosis in a large cohort of middle-aged participants from the Swedish CArdioPulmonary bioImage Study (SCAPIS). We further assessed associations between atherosclerosis-associated gut bacterial species and biomarkers of inflammation and infection, plasma metabolites, and the abundance of corresponding bacterial species in the oral cavity.

## METHODS

### Data Availability

De-identified metagenomic sequencing data can be accessed from the ENA under accession number PRJEB51353 (https://www.ebi.ac.uk/ena/browser/view/prjeb51353). Access to the pseudonymized data requires ethical approval from the Swedish Ethical Review Board and approval from the SCAPIS Data access board (https://www.scapis.org/data-access/). The source code and the summary data underlying all figures used to generate the results for the analysis are available at https://github.com/MolEpicUU/GUTSY_CACS.

### Study Design and Participants

The study design is illustrated in **Figure 1**. The primary data source was SCAPIS, a population-based study focusing on cardiovascular and respiratory disease in 30154 subjects 50–64 years of age from six sites in Sweden. ^13^ Fecal metagenomics data was available for 4839 participants from Uppsala and 4980 from Malmö after removal of 10 samples for failing in the quality control. After excluding subjects with missing information on country of birth (n=39), coronary artery calcium score (CACS) (n=356) or who had self-reported CVD (n=451) including atherosclerotic CVD (n=119), 4541 participants from Uppsala and 4432 from Malmö remained.

**Figure 1.**
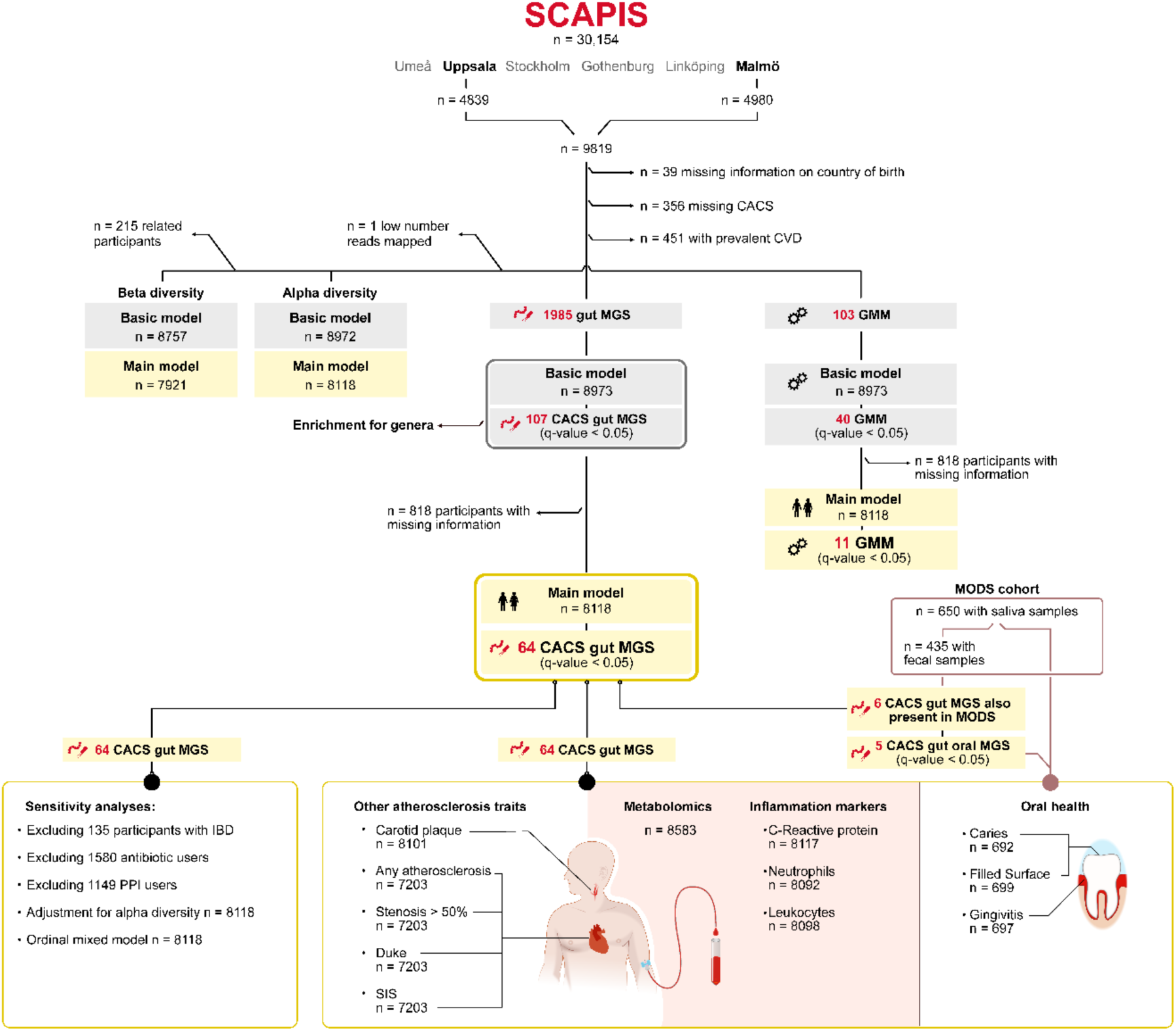
Flowchart of the overall design of the study.

The association between gut species and their counterparts in saliva samples and with oral health was investigated in participants of the Malmö Offspring Dental Study (MODS, n=831, mean age 52.9 years), a substudy of the family-based Malmö Offspring Study (MOS). ^10,14^ Salivary metagenomics, oral examination data, and complete data on age and sex were available for 650 subjects; informative fecal metagenomics data from MOS were available for 435 of those subjects.

All participants gave written informed consent. The study was conducted in accordance with the Declaration of Helsinki and was approved by the regional ethics committees (SCAPIS: Dnr 2010-228-31M and Dnr 2018/315; MODS: Dnr 2013/761; and MOS: Dnr 2012/594).

### Atherosclerosis Measurements

Cardiac imaging in SCAPIS included coronary computed tomography angiography (CCTA) as described previously. ^15^ Briefly, computed tomography was performed with a dedicated dual-source scanner and a Stellar Detector (Somatom Definition Flash, Siemens Medical Solutions). Non-contrast images for calcium scoring were obtained from all subjects, followed by contrast-enhanced images from subjects without contraindications. CACS was determined using the syngo.via calcium scoring software (Volume Wizard; Siemens). The area of calcification was summed for the whole coronary artery tree to a CACS according to Agatston. ^16^ Radiologists inspected the images for any coronary atherosclerosis, segment involvement score (SIS), Duke prognostic coronary artery disease index modified for SCAPIS, ^15^ and occlusive atherosclerosis (≥50% stenosis in at least one vessel) (see **Supplemental Methods)**. We included individuals with valid readings in at least four proximal segments. Carotid atherosclerosis was categorized as absent, unilateral, or bilateral^13^ from two-dimensional gray-scale ultrasound images obtained with a Siemens Acuson S2000 ultrasound scanner and a 9L4 linear transducer (both from Siemens, Germany) using a standardized protocol and the Mannheim consensus. ^17^

### Metagenomics

DNA was extracted from fecal samples (SCAPIS and MOS) and saliva samples (MODS) at Clinical Microbiomics (Copenhagen, Denmark). The analytical pipeline is described in detail in **Supplemental Methods**. In brief, libraries of fragmented DNA were sequenced with the Illumina Novaseq 6000 platform. This technique generated an average of 26.3 million read pairs per sample in SCAPIS-Malmö and MOS, 25.3 million read pairs in SCAPIS-Uppsala, and 26.3 million read pairs in MODS. Non-host reads and data from these and other cohorts were used to build separate nonredundant gene catalogues for fecal and saliva samples. Metagenomic species and corresponding signature gene were identified. ^18^ In all three datasets, species abundance was estimated by mapping reads to the signature gene sets and normalization for effective gene length. The taxonomic information for the two catalogues was mapped with the NCBI RefSeq database. ^19^

Alpha (Shannon diversity index, inverse Simpson index, and Chao1) and beta diversity (Bray-Curtis dissimilarity) were estimated with the R v4.1.1 package *vegan* v2.5-7 using rarefied data.^20^ Rarefied data were also used to define individuals with above/less than median abundance of each species. The functional potential profile of the gut microbiota was determined by assigning genes to gut metabolic modules (GMM),^20^ which includes 103 metabolic pathways that represent cellular enzymatic processes. The GMM abundance was estimated using the *Omixer-RPM* v0.3.226 R package, with a minimum module coverage threshold set at 66.6%. Before conducting statistical analysis, the abundance data for each species in all analytical datasets and for each GMM were transformed with the formula ln(x+1), where x denotes the relative abundance of each species or GMM, followed by z-transformation to set the mean to 0 and SD to 1.

### Clinical Chemistry, Hematology and Metabolomics

Venous plasma samples were analyzed for high-density lipoprotein (HDL) and low-density lipoprotein (LDL) cholesterol, triglycerides, and high-sensitivity C-reactive protein (hsCRP). Blood cells were counted by standard methods. CACS-associated species-plasma metabolite associations in SCAPIS with q-value <0.05 were accessed from the GUTSY atlas (https://gutsyatlas.serve.scilifelab.se/) from Dekkers et al. ^21^

### Oral Health Phenotypes

Dental examinations of MODS participants were done by five dentists. Both surfaces with caries and fillings were recorded on all teeth, counting four to five surfaces per tooth. Surfaces with caries were detected using standard clinical criteria aided by mirror, probe (Hu-Friedy EXD57), and bite-wing radiographs combining initial and manifest lesions. Filled surfaces included both fillings and crowns. Gingival inflammation was recorded as percentage of bleeding on probing, excluding wisdom teeth, and counting six surfaces per tooth.

### Other Phenotypes

In SCAPIS and MOS, data on country of birth, smoking, physical activity, and diet were collected with validated standardized questionnaires. Blood pressure and body mass index (BMI) were recorded. Data on medications were acquired from the drug prescription register, self-report, plasma measurements, and other sources. Relatedness in SCAPIS was based on kinship analysis of genotype data, assigning first-degree relatives individuals to the same family, and relatedness in MODS/MOS was based on family information from the study design phase. Collected phenotypes are described in detail in **Supplemental Methods**.

### Statistical Analysis

#### Association between Gut Microbiota Diversity and Atherosclerosis

The association of alpha diversity with coronary atherosclerosis was assessed by linear mixed models with ln(CACS+1) as the dependent and alpha diversity indices as the independent variables. To address the potential influence of family relatedness, the first degree-relatedness among 423 participants from 208 families was accounted for with a random effect variable by using the *lmerTest* v3.1-3 R package. Furthermore, two sets of covariates were modeled as fixed effects. The basic model was adjusted for age, sex, country of birth, study center, and metagenomics extraction plate. The main model was further adjusted for smoking; physical activity; energy-adjusted carbohydrate, protein, and fiber intake; systolic and diastolic blood pressure; total cholesterol; high-and low-density lipoprotein cholesterol; ln(triglycerides); diabetes; BMI; and self-reported medication for dyslipidemia, hypertension, and diabetes. Correlation of covariates and alpha diversity measurements are reported in **Figure S1**.

Beta diversity was assessed by a distance-based multivariate analysis of variance of ln(CACS+1) as the independent variable using the *dmanova* function in the *GUniFrac* v1.3 package. These analyses were adjusted for the same two set of covariates as used in the alpha diversity analyses. However, in contrast to the alpha diversity analyses, only one participant from each family cluster was included, rather than by treating family relatedness as a random effect variable.

#### Association of Species and Bacterial Functions with CACS

Extensive simulations were conducted before the analysis and supported the use of linear regression to minimize false-positive findings (**Supplemental Methods, Figure S2**). Hence, a series of linear mixed multivariable regressions were performed with the ln(CACS+1) as the dependent variable and the abundance of each species and GMM as the independent variable. The models were fitted for each species and each GMM separately; covariates from the basic model were used as fixed effects and family relatedness as random effects. Multiple testing was accounted for using the Benjamini-Hochberg False Discovery Rate (FDR) at 5%.^22^ A Taxon Set Enrichment Analysis^23^ based on the *fgsea* v1.18.0 R package based on *P* value ranking separately for positive and negative regression coefficients was used to determine whether certain genera were enriched.

Species with a q-value <0.05 in the basic model were assessed adjusting for main model covariates, again accounting for multiple testing. To determine whether associations were caused by influential observations, unscaled dfbeta values were calculated using the *influence* and *dfbetas* R functions. The linear regression estimates were deemed unreliable if the exclusion of the observations with highest absolute dfbeta value resulted in a change of the direction of the regression coefficient or in a *P*≥0.05. Findings not caused by an influential observation are referred to as “CACS-associated species”. Effect estimate modification by sex was assessed for CACS-associated species by entering an interaction term between sex and each of the fixed effect variables in the model and extracting the interaction coefficient of sex with species.

#### Sensitivity Analyses

In three separate analyses, we excluded three groups of participants: those in whom proton-pump inhibitors were measured in plasma, those with inflammatory bowel disease, and those treated with antibiotics during the year before the baseline visit. Furthermore, the analyses for CACS-associated species were repeated with the Shannon diversity index as a covariate to test if findings were driven by global changes in the gut microbiota composition rather than change in individual species. Lastly, the associations from the main model were tested in an ordinal mixed model using CACS categories as the outcome (CACS: 0; 1-100; 101-400; >400) using Stata v15.

#### Association between CACS-Associated Species and CCTA-Based Measurements and Carotid Plaques

We assessed the association of CACS-associated species with presence of any coronary atherosclerotic plaque or coronary stenosis >50% in at least one vessel in a series of logistic mixed regressions. The association between CACS-associated species and SIS, modified Duke index, and number of carotid vessels with atherosclerosis plaques were assessed with ordinal mixed regression models. Categories 4, 5 and 6 of the modified Duke index were merged due to low number of observations. The logistic and ordinal mixed models were fitted using Stata v15 and were adjusted for the main model covariates.

#### Clustering of Plasma Metabolites associated with CACS-Associated Species

For each CACS-associated species, the three strongest associations with plasma metabolites were selected based on their p-value. For the resulting unique subset of 63 metabolites hierarchical clustering was performed based on the Euclidian distance on the partial Spearman correlation coefficients.

#### Associations between CACS-Associated Species and Markers of Systemic Inflammation and Infection

A series of linear mixed models was fitted to assess the association between CACS-associated species (independent variable) and ln-transformed hsCRP, and ln-transformed counts of neutrophils and leukocytes, separately, as the dependent variable. The models were adjusted for the same covariates as the main model.

#### Associations between Gut and Oral CACS-Associated Species and between Oral CACS-Associated Species and Oral Health Phenotypes

To investigate the association between CACS-associated species in the gut and the corresponding species in saliva, MOS/MODS data was analyzed with a series of partial Spearman correlations from a mixed model based on rank-transformed data (details in **Supplemental Methods**). The models were adjusted for age, sex, and metagenomics extraction plate as fixed effects and family relatedness as a random effect. The oral species associated (q-value <0.05) with the corresponding CACS-associated gut species were assessed for association with three oral health phenotypes: filled surfaces (split by deciles in 10 categories), caries, and gingival inflammation. Filled surfaces and caries were assessed using ordinal mixed regressions using the *Ordinal* v2019.12-10 R package, while gingival inflammation was assessed with a linear mixed model. These regression models were adjusted for age, sex, smoking, education, Silness-Löe plaque index, as a marker of oral hygiene, and the activity in the hour immediately before the dental examination (e.g., eating, brushing teeth, smoking) as fixed effects and family relatedness as a random effect.

## RESULTS

### Gut Microbiota Composition and Richness Are Associated with Subclinical Atherosclerosis and Attenuated by Adjustment for Lifestyle Factors, Diet and Medication

The study included 8973 subjects with no self-reported history of CVD. Among them, 40.3% had measurable coronary artery calcification and 5.4% manifested at least one stenosis with >50% occlusion (**Table 1**, **Table S1**). The Shannon diversity index—a measure of overall species richness and evenness within each sample—was inversely associated with CACS (n=8972, β=-0.17, *P*=5.1×10^-4^) in the basic model. The basic model was adjusted for age, sex, country of birth, and technical variables as fixed effects, and family relatedness as a random effect. However, the association was attenuated and non-significant in the main model with more extensive adjustment (n=8118, β=-0.03, *P*=0.53). The covariates primarily attenuating the association were triglycerides and BMI, followed by medication, blood pressure traits, smoking, HDL cholesterol, and physical activity (Figure S3). Alternative alpha diversity indices were similarly associated with CACS (**Figure 2a**). Beta diversity—a measure of the amount of variation in species composition—differed among CACS values in both models, although the main model was attenuated (r^2^basic=0.0005; *P*basic = 2×10^-25^ vs r^2^main=0.0002; *P*main=0.009) mainly by the same covariates that attenuated the relationship between alpha diversity and CACS (**Figure 2b, Figure S3)**. These models were not adjusted for family relatedness, but only one of the participants in each family cluster was included instead (nbasic= 8757, nmain= 7921). These findings encouraged us to investigate species-specific associations.

**Table 1.**
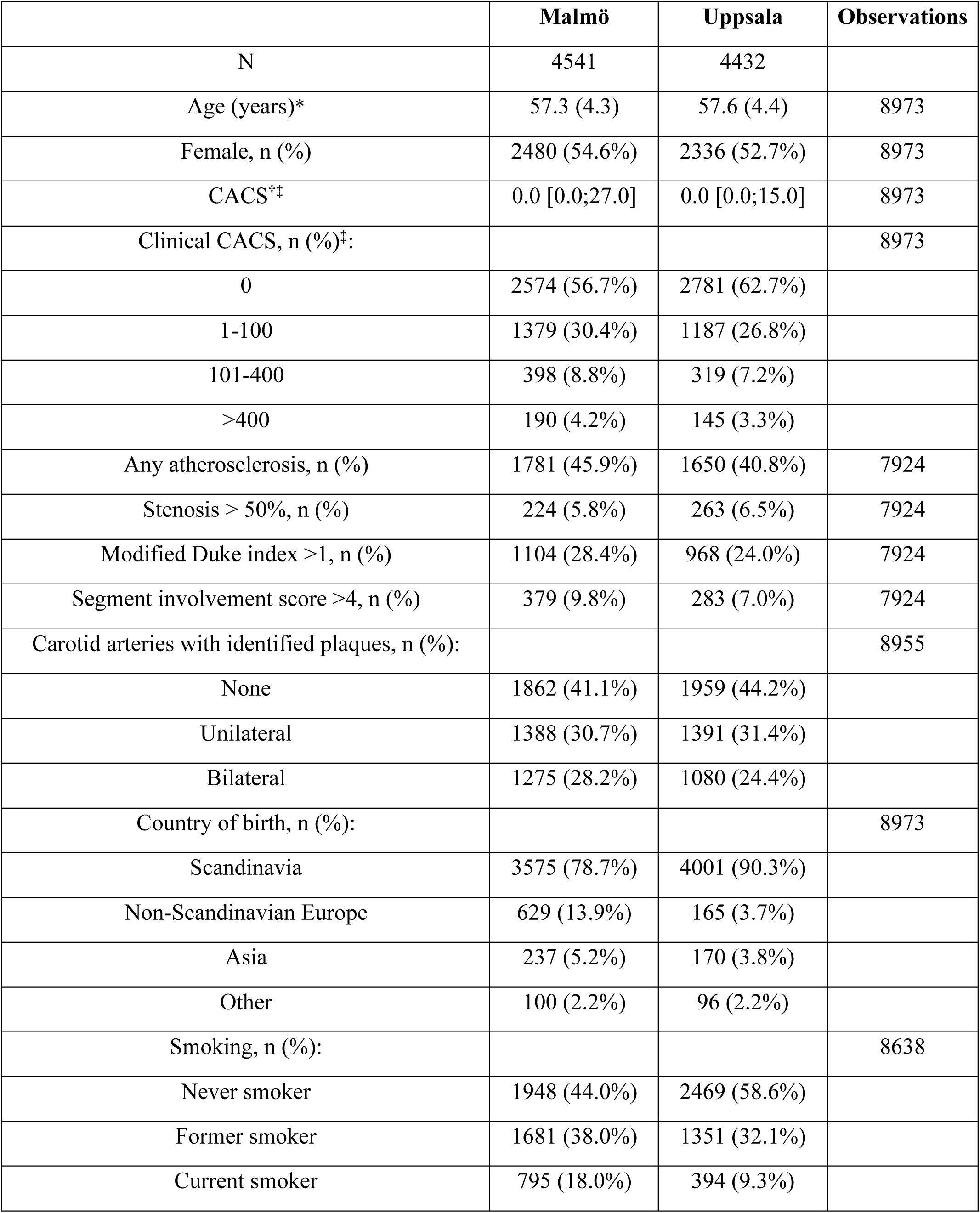
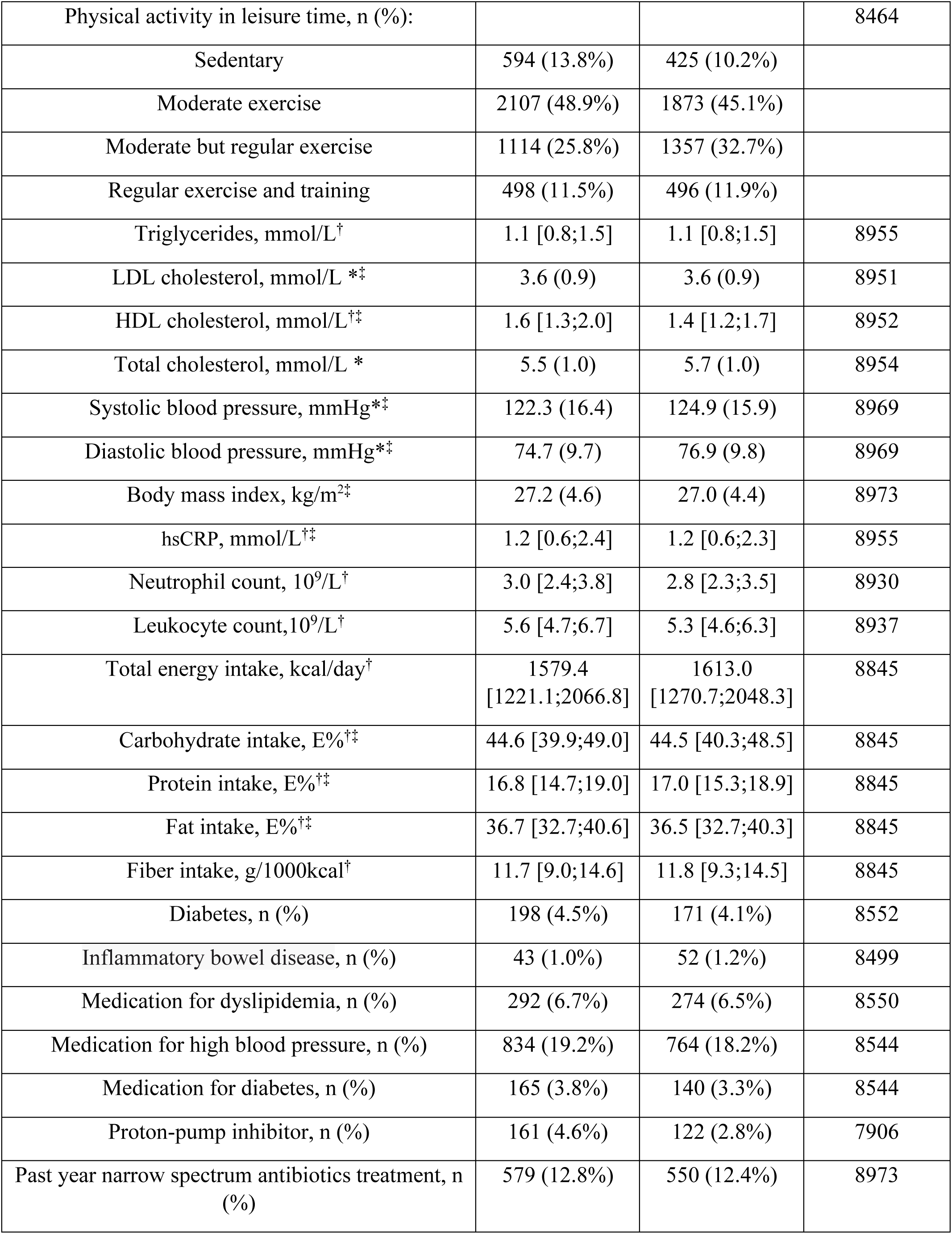
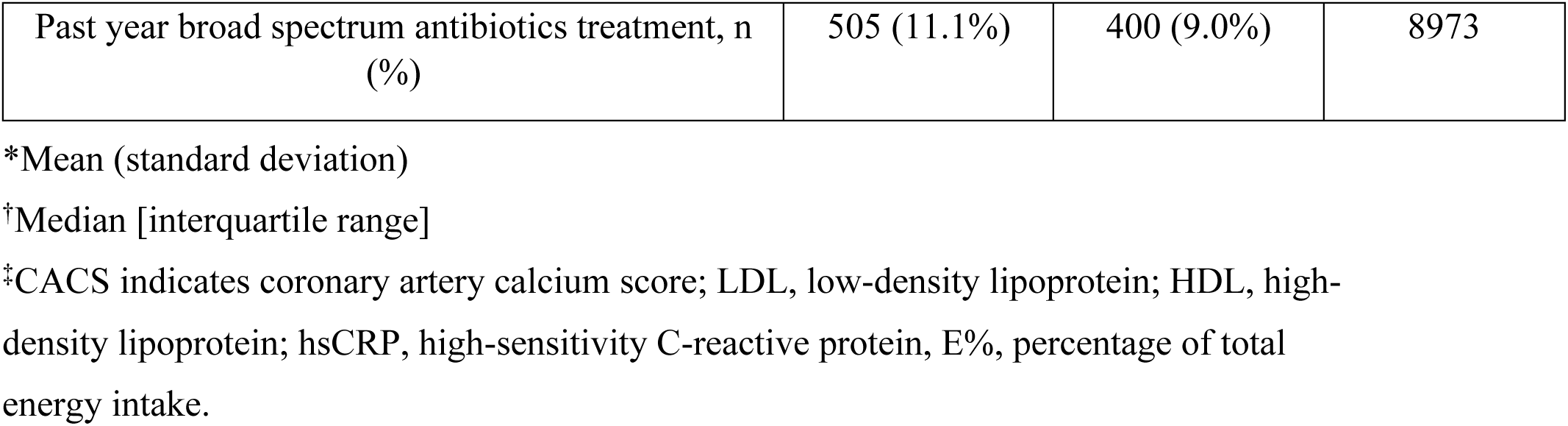
Descriptive characteristics of participants in the SCAPIS study with metagenomics data and no previous CVD event

|  | **Malmö** | **Uppsala** | **Observations** |
| --- | --- | --- | --- |
| N | 4541 | 4432 |  |
| Age (years)* | 57.3 (4.3) | 57.6 (4.4) | 8973 |
| Female, n (%) | 2480 (54.6%) | 2336 (52.7%) | 8973 |
| CACS<sup>†‡</sup> | 0.0 [0.0;27.0] | 0.0 [0.0;15.0] | 8973 |
| Clinical CACS, n (%)<sup>‡</sup>: |  |  | 8973 |
| 0 | 2574 (56.7%) | 2781 (62.7%) |  |
| 1-100 | 1379 (30.4%) | 1187 (26.8%) |  |
| 101-400 | 398 (8.8%) | 319 (7.2%) |  |
| >400 | 190 (4.2%) | 145 (3.3%) |  |
| Any atherosclerosis, n (%) | 1781 (45.9%) | 1650 (40.8%) | 7924 |
| Stenosis > 50%, n (%) | 224 (5.8%) | 263 (6.5%) | 7924 |
| Modified Duke index >1, n (%) | 1104 (28.4%) | 968 (24.0%) | 7924 |
| Segment involvement score >4, n (%) | 379 (9.8%) | 283 (7.0%) | 7924 |
| Carotid arteries with identified plaques, n (%): |  |  | 8955 |
| None | 1862 (41.1%) | 1959 (44.2%) |  |
| Unilateral | 1388 (30.7%) | 1391 (31.4%) |  |
| Bilateral | 1275 (28.2%) | 1080 (24.4%) |  |
| Country of birth, n (%): |  |  | 8973 |
| Scandinavia | 3575 (78.7%) | 4001 (90.3%) |  |
| Non-Scandinavian Europe | 629 (13.9%) | 165 (3.7%) |  |
| Asia | 237 (5.2%) | 170 (3.8%) |  |
| Other | 100 (2.2%) | 96 (2.2%) |  |
| Smoking, n (%): |  |  | 8638 |
| Never smoker | 1948 (44.0%) | 2469 (58.6%) |  |
| Former smoker | 1681 (38.0%) | 1351 (32.1%) |  |
| Current smoker | 795 (18.0%) | 394 (9.3%) |  |
| Physical activity in leisure time, n (%): |  |  | 8464 |
| Sedentary | 594 (13.8%) | 425 (10.2%) |  |
| Moderate exercise | 2107 (48.9%) | 1873 (45.1%) |  |
| Moderate but regular exercise | 1114 (25.8%) | 1357 (32.7%) |  |
| Regular exercise and training | 498 (11.5%) | 496 (11.9%) |  |
| Triglycerides, mmol/L<sup>†</sup> | 1.1 [0.8;1.5] | 1.1 [0.8;1.5] | 8955 |
| LDL cholesterol, mmol/L *<sup>‡</sup> | 3.6 (0.9) | 3.6 (0.9) | 8951 |
| HDL cholesterol, mmol/L<sup>†‡</sup> | 1.6 [1.3;2.0] | 1.4 [1.2;1.7] | 8952 |
| Total cholesterol, mmol/L * | 5.5 (1.0) | 5.7 (1.0) | 8954 |
| Systolic blood pressure, mmHg*<sup>‡</sup> | 122.3 (16.4) | 124.9 (15.9) | 8969 |
| Diastolic blood pressure, mmHg*<sup>‡</sup> | 74.7 (9.7) | 76.9 (9.8) | 8969 |
| Body mass index, kg/m<sup>2‡</sup> | 27.2 (4.6) | 27.0 (4.4) | 8973 |
| hsCRP, mmol/L<sup>†‡</sup> | 1.2 [0.6;2.4] | 1.2 [0.6;2.3] | 8955 |
| Neutrophil count, 10<sup>9</sup>/L<sup>†</sup> | 3.0 [2.4;3.8] | 2.8 [2.3;3.5] | 8930 |
| Leukocyte count,10<sup>9</sup>/L<sup>†</sup> | 5.6 [4.7;6.7] | 5.3 [4.6;6.3] | 8937 |
| Total energy intake, kcal/day<sup>†</sup> | 1579.4<br>[1221.1;2066.8] | 1613.0<br>[1270.7;2048.3] | 8845 |
| Carbohydrate intake, E%<sup>†‡</sup> | 44.6 [39.9;49.0] | 44.5 [40.3;48.5] | 8845 |
| Protein intake, E%<sup>†‡</sup> | 16.8 [14.7;19.0] | 17.0 [15.3;18.9] | 8845 |
| Fat intake, E%<sup>†‡</sup> | 36.7 [32.7;40.6] | 36.5 [32.7;40.3] | 8845 |
| Fiber intake, g/1000kcal<sup>†</sup> | 11.7 [9.0;14.6] | 11.8 [9.3;14.5] | 8845 |
| Diabetes, n (%) | 198 (4.5%) | 171 (4.1%) | 8552 |
| Inflammatory bowel disease, n (%) | 43 (1.0%) | 52 (1.2%) | 8499 |
| Medication for dyslipidemia, n (%) | 292 (6.7%) | 274 (6.5%) | 8550 |
| Medication for high blood pressure, n (%) | 834 (19.2%) | 764 (18.2%) | 8544 |
| Medication for diabetes, n (%) | 165 (3.8%) | 140 (3.3%) | 8544 |
| Proton-pump inhibitor, n (%) | 161 (4.6%) | 122 (2.8%) | 7906 |
| Past year narrow spectrum antibiotics treatment, n (%) | 579 (12.8%) | 550 (12.4%) | 8973 |
| Past year broad spectrum antibiotics treatment, n (%) | 505 (11.1%) | 400 (9.0%) | 8973 |

\*Mean (standard deviation)
†Median [interquartile range]
‡CACS indicates coronary artery calcium score; LDL, low-density lipoprotein; HDL, high- density lipoprotein; hsCRP, high-sensitivity C-reactive protein, E%, percentage of total energy intake.

**Figure 2.**
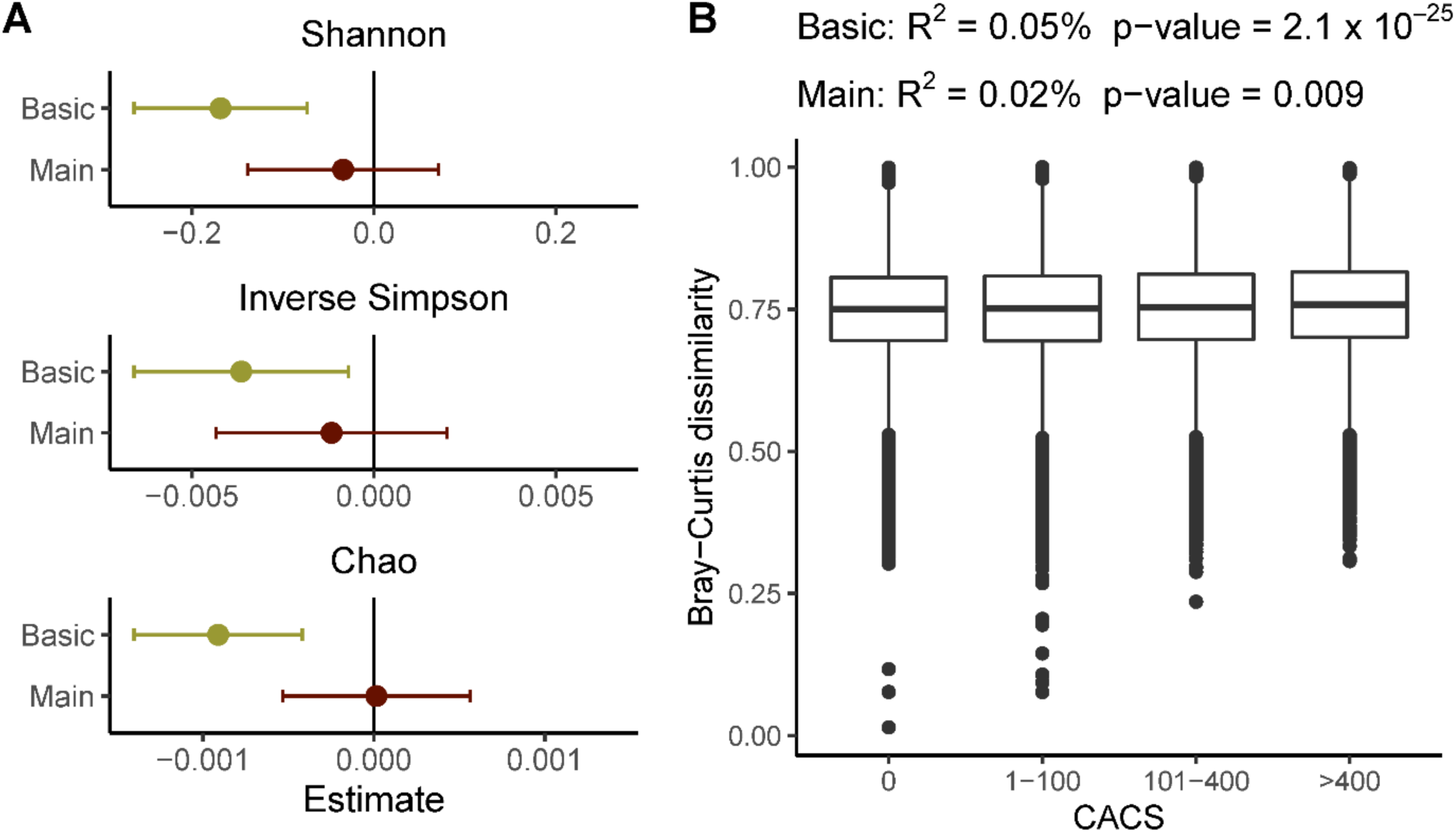
Association of alpha and beta diversity with CACS. The association of alpha diversity (a) with coronary atherosclerosis was assessed by fitting linear mixed models with CACS as the outcome and the alpha diversity indices as the exposure. The basic model (n=8972) was adjusted for age, sex, country of birth, and metagenomics extraction plate as fixed effects and family relatedness as random effect. The main model (n=8118) was further adjusted for, smoking, physical activity, energy-adjusted intake of carbohydrate, protein, and fiber, systolic blood pressure (SBP), and diastolic blood pressure (DBP), total cholesterol, HDL and LDL cholesterol, triglycerides, BMI, diabetes, and self-reported medication for dyslipidemia, hypertension, and diabetes as fixed effects. For beta diversity (b), a distance-based multivariate analysis of variance of CACS as the independent variable was applied and the same fixed effect adjustments were made without the random effect adjustment. In the basic (n=8757) and the full model (n=7921) one participants for each family relatedness cluster was removed. Box plots represent the Bray-Curtis dissimilarity between participants with CACS=0 and the other CACS groups. For the CACS=0, the boxplot represents the within group dissimilarity.

### Species Associated with Subclinical Atherosclerosis Are Enriched in Species from ***Streptococcus* and *Oscillibacter* Genera**

The taxonomic profiling of fecal samples identified the prevalence of 1985 species, with 411 species having a prevalence <1%. The average number of species per sample was 325, with a range of 11 to 696. The dominant phyla were Firmicutes (72%) and Bacteroidetes (20%) (**Figure 3**). In the basic model, including 8973 individuals, the relative abundance of 78 species (73 of which had a prevalence ≥1%) showed a positive association with CACS, while 29 species (all of which had a prevalence ≥1%) were negatively associated. (**Table S2**). Taxon-set enrichment analysis revealed an overrepresentation of the *Streptococcus* and *Oscillibacter* genera in the positive CACS associations (**Table S3**).

**Figure 3.**
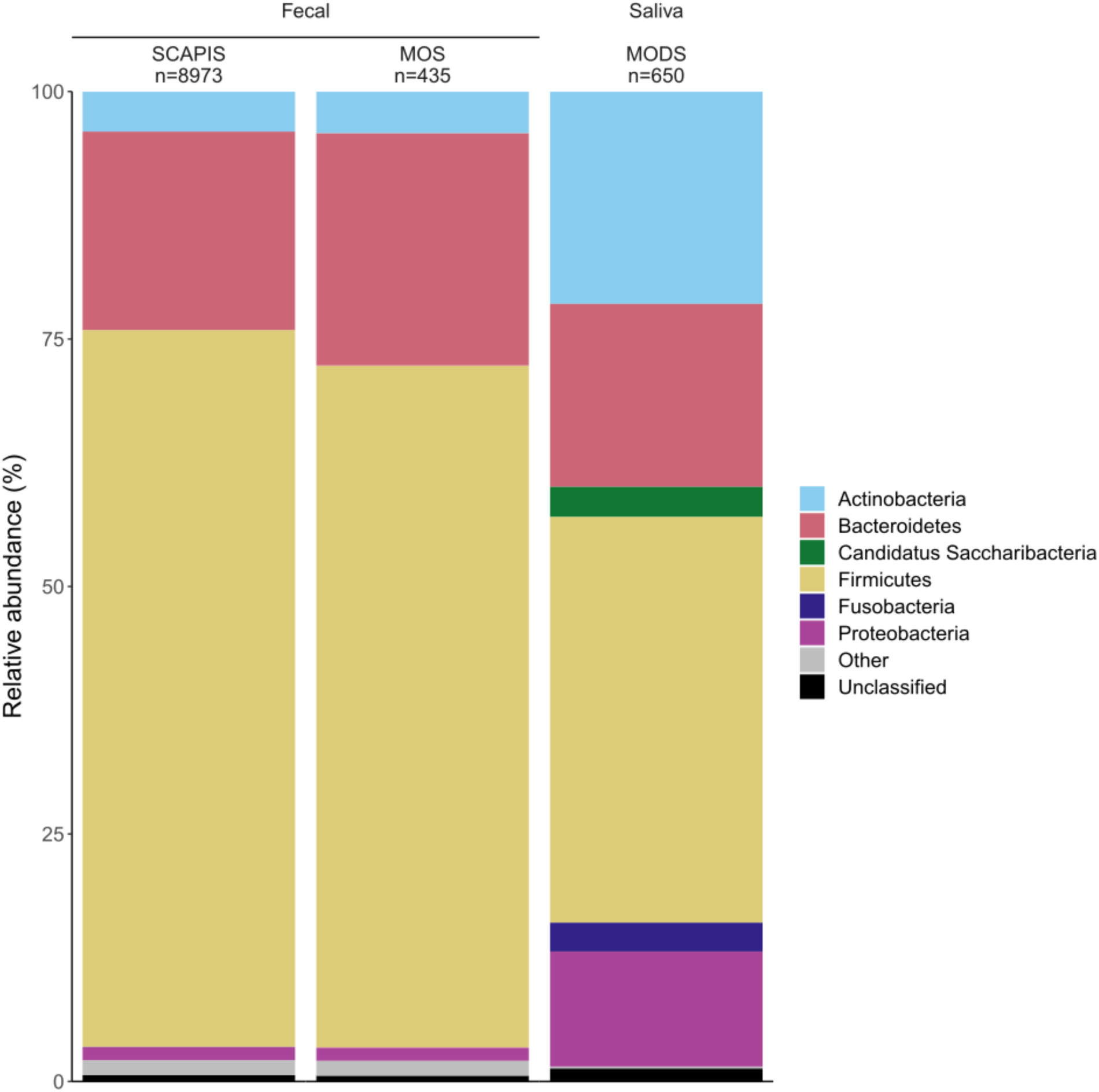
Phylum-level taxonomical profiling of the fecal and saliva samples across the three data sets.

### Sixty-four Species Were Associated with CACS Independent of Cardiovascular Risk Factors

In the main model, which included 8118 individuals with complete data on covariates, 67 species remained associated with CACS (54 positively and 13 negatively) (**Figure 4**). Covariates included age, sex, country of birth, metagenomics extraction plate, smoking, physical activity, energy-adjusted carbohydrate, protein, fiber intake, blood pressure, lipids, diabetes, BMI, self-reported cardiovascular medication, and family relatedness. However, the associations of *Peptoniphilus harei*, *Muribaculaceae* sp. (MGS identifier: HG3A.1967) and *Eubacteriales* sp. (HG3A.0270) were deemed non-robust, as the removal of single influential observations abolished the association (**Table S4**). Among the remaining 64 CACS-associated species, 51 had a positive association with CACS, while 13 had negative associations (**Figure 4**, **Figure 5**). The abundance of bacteria across CACS groups and the previously excluded SCAPIS participants with self-reported atherosclerotic CVD is shown in **Figure S4**. The lowest *P* values were observed for *Streptococcus anginosus*, *S. oralis* subsp. *oralis*, *Escherichia coli*, *Eubacteriales* sp. (HG3A.1354) and *Intestinimonas* sp. (HG3A.1149), all positively associated with CACS. Other associated species included *S. parasanguinis*, *S. gordonii*, and *S. agalactiae*. Comparison of the main clinical characteristics in participants with high and low abundance of CACS-associated species (cut-off median) showed that those with higher abundance of the two most strongly associated species, *S. anginosus,* and *S. oralis* subsp. *oralis* had in general more cardiovascular risk factors (**Table S5**). Furthermore, there were indications that the associations between *S. agalactiae*, *Rothia mucilaginosa*, two *Eubacteriales* spp. (HG3A.0511 and HG3A.0854), and an *Oscillibacter* sp. (HG3A.0243) and CACS were modified by sex (interaction *P*<0.05, **Table S4**). After adjusting for multiple testing the associations were no longer significant.

**Figure 4.**
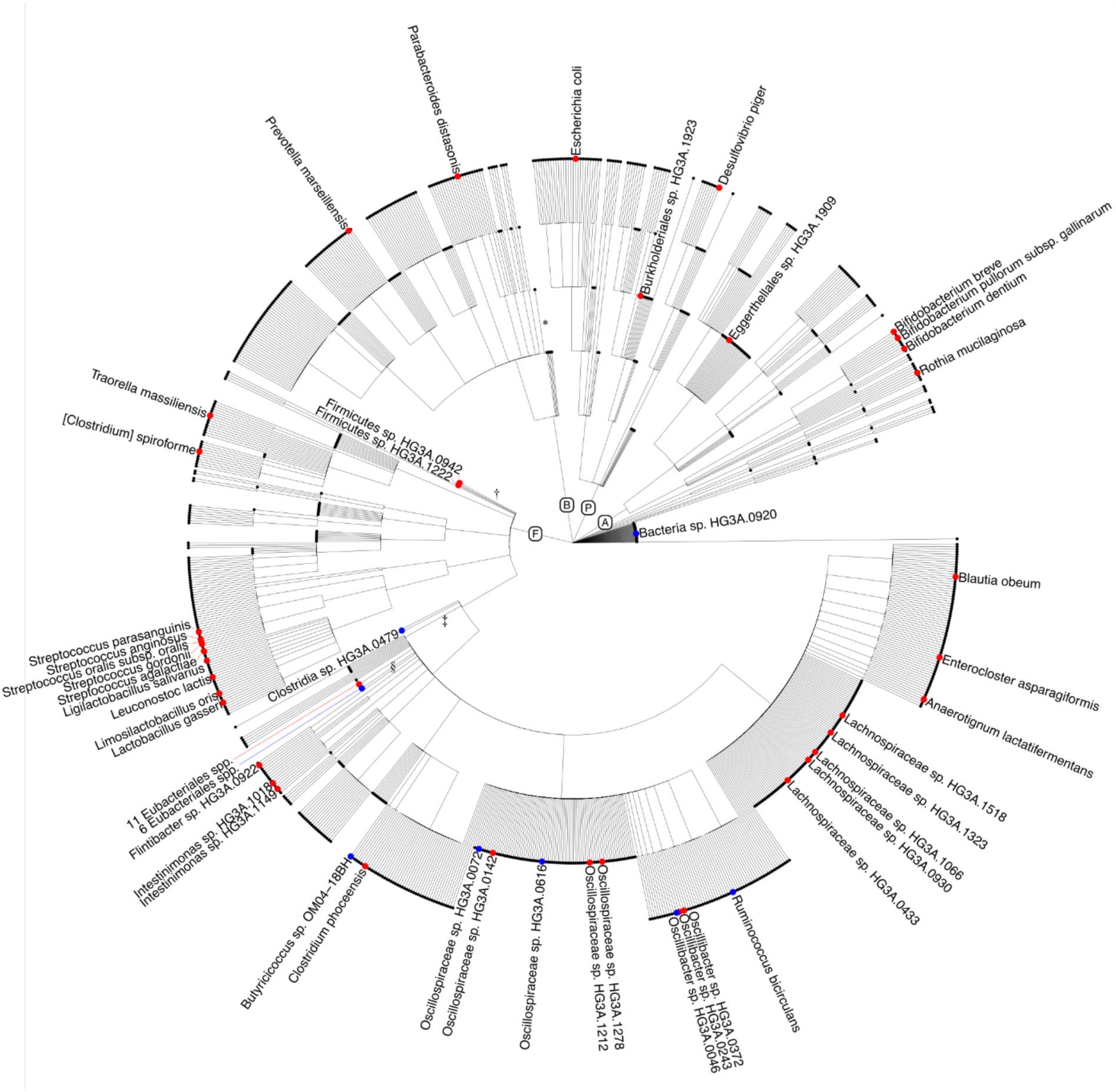
Cladogram of species investigated in the study. Circles represent metagenomic species annotated to the lowest taxonomic level. Dots indicate the species positively (red) and negatively (blue) associated with CACS in the main model (false discovery rate <5%). A: *Actinobacteria*; P: *Proteobacteria*; B: *Bacteroides*; F: *Firmicutes*. The following unclassified non-CACS-associated metagenomics species were collapsed: *99 species with lowest annotation to the order *Bacteroidales*, ^†^95 species with lowest annotation to phylum *Firmicutes*, ^‡^163 species with lowest annotation to the class *Clostridia*, and ^§^651 species with lowest annotation to the order *Eubacteriales*.

**Figure 5.**
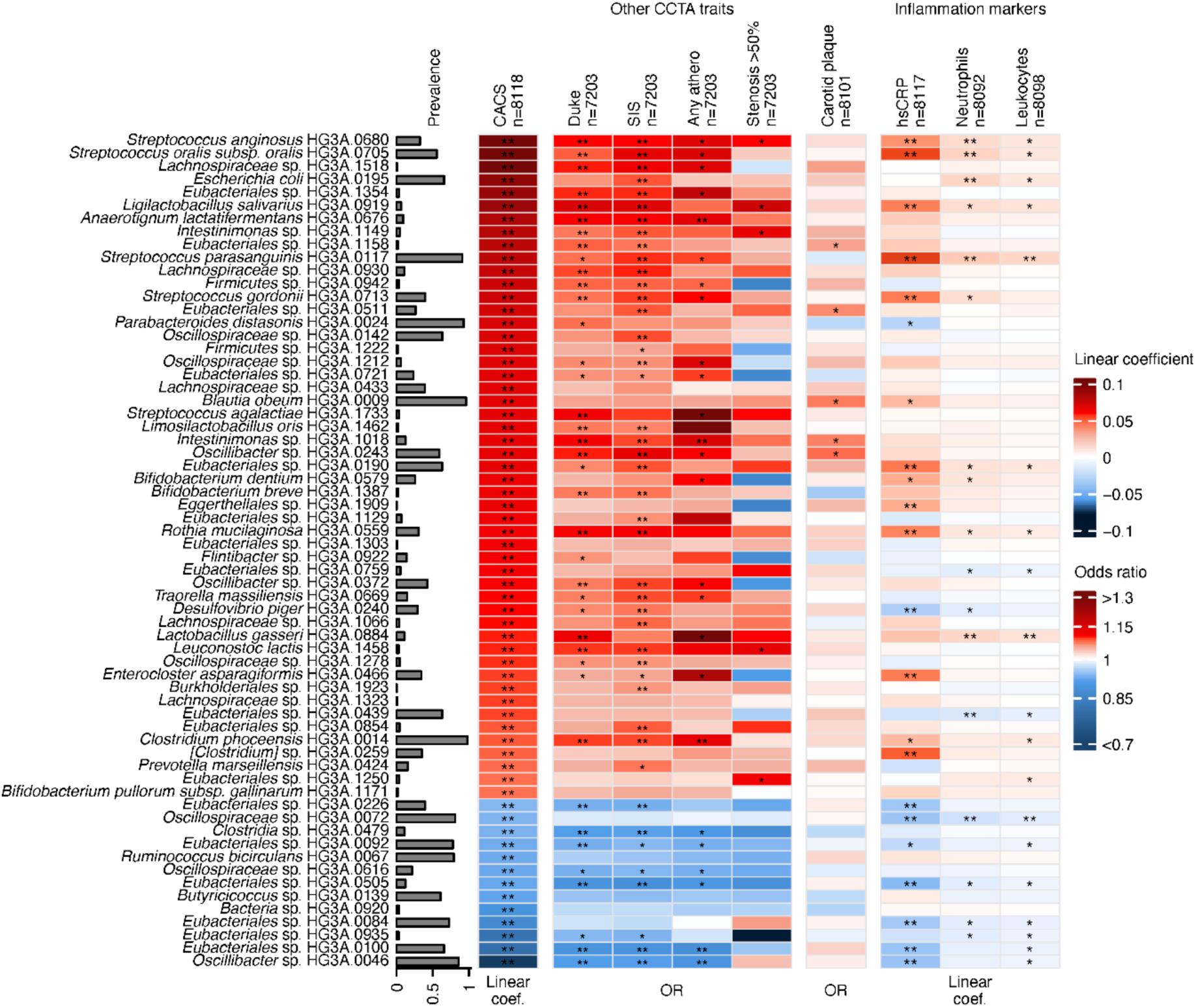
Associations between CACS-related gut species and alternate measurements of atherosclerosis and markers of inflammation and infection. Heatmap showing the associations of the 64 coronary artery calcium score (CACS)-associated species (False Discovery Rate (FDR) q-value<0.05) with CACS, other coronary computed tomography angiography atherosclerosis traits, carotid plaque, and inflammation biomarkers. Associations with CACS and with inflammatory markers hsCRP (high-sensitivity C-reactive protein), neutrophil and leukocyte counts are presented as linear regression coefficients after adjustment for the main model covariates (age, sex, country of birth, metagenomics extraction plate, smoking, physical activity, energy-adjusted carbohydrate, protein and fiber intake, systolic and diastolic blood pressure, high-density lipoprotein (HDL), low-density lipoprotein (LDL), and total cholesterol, triglycerides, body mass index (BMI), diabetes, and self-reported medication for dyslipidemia, hypertension, and diabetes as fixed effects and first-degree family relatedness as a random effect). Associations with atherosclerosis traits (i.e., modified Duke index, segment involvement score (SIS), any atherosclerosis, and ≥50% stenosis) and presence of carotid plaques are presented as odds ratio (OR) after adjustment for main model covariates. Associations with FDR q-value <0.05 are marked with two asterisks (**) and those with P<0.05 are marked with one asterisk (*).

### Sensitivity Analyses

The exclusion of individuals who used proton-pump inhibitors, had inflammatory bowel disease, or were treated with antibiotics in the previous 12 months before the baseline visit, as well as adjusting for Shannon diversity index showed largely similar effect estimates as the main model (**Figure S5**). Applying ordinal mixed models showed consistent effect direction and *P*<0.05 for 61 out of 64 species (**Table S4**). Overall, these findings indicate that the associations identified in the main analysis were reliable.

### Gut Metabolic Modules Related to Dissimilatory Nitrate Reduction, Anaerobic Fatty Acid Beta-oxidation and Amino Acid Degradation are Positively Associated with Subclinical Atherosclerosis

In the basic model (n = 8973), 36 GMMs were positively associated while 4 had a negative association with CACS (**Table S6**). In the main model (n=8118), 11 GMMs were found to be positively associated with CACS, and none were negatively associated with CACS (**Table S7**). None of these associations were driven by an influential observation. These 11 GMMs are involved in various metabolic pathways, including amino acid degradation (e.g., asparagine, proline, and valine degradation), anaerobic fatty acid beta-oxidation, enzymatic reactions on lactate consumption, propionate production, and nitrogen metabolism.

### Other Atherosclerosis Measurements

Next, we assessed the association between CACS-associated species and four CCTA measurements (presence of coronary atherosclerosis plaque, coronary stenosis >50%, modified Duke index, and SIS) and the number of carotid vessels with atherosclerotic plaque adjusted for the covariates in the main model. Twenty-five CACS-associated species were also associated with any coronary atherosclerotic plaque, 5 with coronary stenosis >50%, 39 with the modified Duke index, 44 with the SIS, and 5 with carotid plaque (*P*<0.05), all in the same direction as with CACS (**Figure 5**). The correlation among atherosclerosis measurements is shown in **Figure S6**.

### CACS-Associated Species are Linked to Numerous Plasma Metabolites and Clustered into Distinct Groups Based on their Strongest Associations

We assessed whether the 60 CACS-associated species available in the GUTSY Atlas^21^ describing species-metabolite associations were associated with plasma metabolites. All 60 species had at least one association with a plasma metabolite and there were 2500 associations (1478 positive and 1022 negative) between CACS-associated species and metabolites in total (**Table S8**). Based on their strongest associations with metabolites the species clustered in distinct groups, with one group containing species commonly found in the oral microbiome^24^ (e.g., all CACS-associated *Streptococcus* spp., *R. mucilaginosa, Bifidobacterium dentium*, and *Ligilactobacillus salivarius*). Amongst others, this group was negatively associated with a set of uncharacterized molecules, and with indole propionate and positively associated with secondary bile acids and imidazole propionate. The remaining species were dominated by *Eubacteriales* sp., and included both CACS-positive and negatively associated species and had an inverse metabolic pattern compared to the oral microbiome set, with the exception of tobacco-related metabolites, which were positively associated with CACS-positively associated species and negatively associated with CACS-negatively associated species (**Figure S7**).

### Oral Species in the Gut Microbiota are Associated with Markers of Systemic Inflammation and Infection

We assessed associations between CACS-associated species and plasma hsCRP (n=8117) and blood counts of leukocytes (n=8098), and neutrophils (n=8092) in linear mixed models. Of 54 species positively associated with CACS, 13 were also positively associated with hsCRP, 10 with leukocyte counts, and 11 with neutrophil counts (*P*<0.05). Species associated with all three markers were dominated by species common in the oral microbiome^24^ including the streptococci species most strongly associated with CACS: *S. anginosus*, *S. oralis* subsp*. oralis*, and *S. parasanguinis.* Among the 13 species negatively associated with CACS, 6 were negatively associated with hsCRP, 6 with leukocyte counts, and 5 with neutrophil counts (**Figure 5**).

### CACS-Associated Species from Fecal Samples Correlate with Corresponding Species in Saliva

Several of the CACS-associated species are normally considered as oral species. ^24^ We therefore investigated the correlation between the abundance of CACS-associated species in the fecal and saliva samples from the 435 participants in MODS (**Table S9**) who underwent a thorough dental examination within 4 to 12 months after fecal sampling in the MOS. Six of the CACS-associated species were identified in the saliva samples, of which five (*S. anginosus, S. parasanguinis*, *S. gordonii*, *R. mucilaginosa*, and *B. dentium*) were positively associated with the abundance of the corresponding species in the fecal sample (rho=0.15–0.46), while 1. *S. oralis* subsp. *oralis* was not (**Table S10**). In up to 639 participants from MODS, three species (*S. anginosus, S. parasanguinis,* and *R. mucilaginosa*) were positively associated with both caries and filled surfaces, and three (*S. anginosus, S. gordonii and B. dentium*) with gingivitis (**Table S11**).

## DISCUSSION

Gut bacteria have been proposed to affect the development and progression of atherosclerosis through infections local or distal to the atherosclerotic plaque or through production of atherogenic metabolites. ^2^ The association of gut microbiota with coronary atherosclerosis has previously been studied only in symptomatic patients, who are often under treatment, resulting in high risk of bias as medication may result in large shifts of the composition. Here, detailed image-based measurements of coronary artery atherosclerosis and deep characterization of the gut microbiome by shotgun metagenomics of individuals without earlier cardiovascular disease in the large population-based SCAPIS cohort were used to identify 64 species associated with CACS independent of risk factors. Several of these species were associated with circulating markers of inflammation and infection, and with the corresponding bacteria species in saliva samples, which were in turn associated with worse oral health. Thus, studies are merited to continue to investigate the role of these 64 species in atherogenesis.

The *Streptococcus* genus was associated with CACS, consistent with observations in previous case-control studies of symptomatic atherosclerotic cardiovascular disease. ^6,7,25^ Specifically, *S. anginosus*, *S. oralis* subsp. *oralis*, *S. parasanguinis*, *S. gordonii*, and *S. agalactiae* were associated with coronary atherosclerosis. Current knowledge of the involvement of these bacteria in cardiovascular pathologies is summarized in **Table S12**. These species all belong to the VGS, ^26^ except for *S. agalactiae* (a β-hemolytic non-VGS). VGS can enter the bloodstream through mucosal barrier injuries from daily dental care activities and dental procedures, and could also pass the gut barrier when injured. ^27^ VGS can infect the valves and the coronary vessels, accounting for 20% of infective endocarditis cases. ^4,9,28^ VGS species initiate and contribute to biofilm formations, which enhance bacterial survival and have been found in atherosclerotic lesions with unclear significance. ^29^ Moreover, in model systems, VGS can invade human aortic endothelial cells and stimulate atherosclerosis-related pro-inflammatory cytokines. ^30^ Animal studies suggest a causal link between *Streptococcus* spp. and atherogenesis. ^31,32^ In our study, the abundance of CACS-associated *Streptococcus* spp. in the gut associated strongly and positively with hsCRP, leukocytosis, and neutrophilia, which could have been triggered by low-grade bacteremia.

The abundance of CACS-associated species might be reduced by antimicrobial treatment. However, clinical trials showed that anti-infective therapies, targeted at chlamydial colonization of the coronary plaques, seemed harmful in patients with established coronary heart disease. ^33^ In the CLARICOR study, mortality was increased in patients with stable coronary artery disease treated with clarithromycin. ^34^ Possible explanations to why these antibiotics were ineffective in reducing coronary events include resistance to antibiotics due to biofilm formation, ^35^ recolonization after antibiotic treatment, the need for treatment earlier in atherosclerosis process, or simply that these bacteria are not causal to atherosclerosis. In the current study, participants with low abundance of CACS-associated *Streptococcus* spp. were not more exposed to antibiotics over the past 12 months than those with higher abundance. We also found that the abundance of *Streptococcus* spp. was associated with coronary artery calcification even after exclusion of participants treated with antibiotics.

We assessed the association of the CACS-associated species with other measures of coronary atherosclerosis from the CCTA as well as with carotid atherosclerosis measured by ultrasound. In general, we found high agreement of associations of species with CACS, modified Duke index, any atherosclerosis, and SIS. However, the associations with having an occlusion >50% of at least one vessel were weaker, perhaps because this was a rather rare phenotype in our cohort, with low power. Likewise, only a few of the species were associated with carotid plaque, perhaps because the measurement of carotid plaque was not very detailed or because associations differ in different vascular beds.

The gut microbiota composition might also contribute to atherogenesis through alteration of the host metabolism. In the current study, all CACS-associated species were associated with at least one plasma metabolite and clustered in distinct groups based on their associations with the metabolome. One group, which contained all the species commonly found in saliva, was positively associated with several microbiota-derived metabolites such as primary and secondary bile acids, and with omeprazole and metformin. Furthermore this group was negatively associated with the microbially-derived tryptophan metabolite indole propionate, a metabolite which has been found inversely associated with atherosclerotic coronary disease in humans and reduced progression of atherosclerosis in mice. ^36^ Bacteria of this group were also found positively associated with imidazole propionate, a microbially-derived metabolite from histidine, reported to impair glucose metabolism. ^37^ Several tobacco metabolites, i.e. 3-hydroxycotinine glucuronide, cotinine N-oxide and norcotinine, were positively associated with species positively associated with CACS and negatively associated with species negatively associated with CACS. Together, these findings suggest an interplay between CACS-associated species and microbial metabolite production, drug intake and smoking behavior.

Another aspect of the microbiome is the functional metabolic potential, which can be summarized into metabolic modules based on known functions of bacterial genes. ^20^ We found eleven modules positively associated with CACS, with the strongest associations noted for dissimilatory nitrate reduction and anaerobic fatty acid beta-oxidation. Dietary nitrate is absorbed in the intestines and excreted back into the gastrointestinal tract by the salivary glands at high concentrations, forming the so-called entero-salivary circulation of nitrate. Denitrifying oral bacteria are important for converting nitrate (NO3^−^) into nitrite (NO2^−^), which can be further converted into nitric oxide (NO) in the acidic pH of the stomach. The generation of NO contributes to the gastric protection by increasing blood flow and mucus thickness. ^38^ The nitrate and nitrite that is not converted to NO in the stomach is absorbed into the bloodstream and tissues, where they might have act as reservoir for NO, which appear to be important for vasodilation under hypoxia^39^ and modulation of mitochondrial respiration. ^40^ However, the dissimilatory nitrate reduction pathway converts nitrate to ammonia, and having an increased activity of this pathway in the colon could potentially inhibit the potential positive cardiovascular effects of nitrate. We found that genes related to anaerobic fermentation of fatty acids were associated with CACS. Such genes are rather uncommon in the gut microbiota and the genetic background of oxidation of fatty acids under anaerobic conditions is not well characterized. We found an association of five amino acid breakdown modules with CACS. This could be driven by differences in diet not captured by our covariates.

Our study has some limitations. First, although our cohort is at least seven times larger than those analyzed previously, few participants had high levels of subclinical atherosclerosis, reducing statistical power. Second, microbial composition can vary extensively throughout the gastrointestinal tract. Fecal samples contain microbial populations from the distal colon, and less so from other sites such as the small intestine. Therefore, we could not identify associations of species that are not well represented in fecal samples. Third, our study does not take into consideration the different interactions among bacterial species such as synergistic effects in the relationship with coronary atherosclerosis. Fourth, the cross-sectional study design limits causal inference. Finally, our data on antibiotic treatment does not capture antibiotics provided in inpatient care; however, in this age group, we do not expect a large number of hospital-treated patients. In future studies, different causal inference methods should be used to determine whether the identified species are causally related to atherosclerosis development.

In conclusion, by combining data from a large population-based cohort study and imaging to evaluate subclinical coronary atherosclerosis, we found that the abundance of several species in the gut were associated with coronary atherosclerosis, biomarkers of inflammation, and their oral counterparts. If causal, these species might contribute to atherogenesis by direct infection or by altering host metabolism. Future studies will show whether these species can be used as potential biomarkers or treatment targets.

## Supplemental Material

Supplemental Methods

Tables S1-S12

Figures S1-S7

## Supporting information

STORMS checklist

## Data Availability

De-identified metagenomic sequencing data can be accessed from the ENA under accession number PRJEB51353 (https://www.ebi.ac.uk/ena/browser/view/prjeb51353). Access to the pseudonymized data requires ethical approval from the Swedish Ethical Review Board and approval from the SCAPIS Data access board (https://www.scapis.org/data-access/).

https://github.com/MolEpicUU/GUTSY_CACS

https://www.ebi.ac.uk/ena/browser/view/prjeb51353

## Non-standard Abbreviation and Acronyms

CACS: Coronary artery calcium score
CCTA: Coronary computed tomography angiography
CVD: Cardiovascular disease
GMM: Gut metabolic module
hsCRP: High-sensitivity C-reactive protein
ln: Natural logarithm
MGS: Metagenomics species
MODS: Malmö Offspring Dental Study
MOS: Malmö Offspring Study
SCAPIS: Swedish CArdioPulmonary bioImage Study
SIS: Segment involvement score
VGS: Viridans streptococci group

## REFERENCES

1. Roth GA, Mensah GA, Johnson CO, Addolorato G, Ammirati E, Baddour LM, Barengo NC, Beaton AZ, Benjamin EJ, Benziger CP, et al. Global Burden of Cardiovascular Diseases and Risk Factors, 1990–2019: Update From the GBD 2019 Study. Journal of the American College of Cardiology. 2020;76:2982–3021. doi: https://doi.org/10.1016/j.jacc.2020.11.010

2. Jonsson AL, Bäckhed F. Role of gut microbiota in atherosclerosis. Nat Rev Cardiol. 2017;14:79–87. doi: 10.1038/nrcardio.2016.183

3. Chakaroun RM, Olsson LM, Backhed F. The potential of tailoring the gut microbiome to prevent and treat cardiometabolic disease. Nat Rev Cardiol. 2023;20:217–235. doi: 10.1038/s41569-022-00771-0

4. Ott SJ, Mokhtari NEE, Musfeldt M, Hellmig S, Freitag S, Rehman A, Kühbacher T, Nikolaus S, Namsolleck P, Blaut M, et al. Detection of diverse bacterial signatures in atherosclerotic lesions of patients with coronary heart disease. Circulation. 2006;113:929–937. doi: 10.1161/CIRCULATIONAHA.105.579979

5. Fromentin S, Forslund SK, Chechi K, Aron-Wisnewsky J, Chakaroun R, Nielsen T, Tremaroli V, Ji B, Prifti E, Myridakis A, et al. Microbiome and metabolome features of the cardiometabolic disease spectrum. Nature Medicine 2022. 2022;37:1–12. doi: 10.1038/s41591-022-01688-4

6. Jie Z, Xia H, Zhong SL, Feng Q, Li S, Liang S, Zhong H, Liu Z, Gao Y, Zhao H, et al. The gut microbiome in atherosclerotic cardiovascular disease. Nature Communications. 2017;8:845. doi: 10.1038/s41467-017-00900-1

7. Liu H, Chen X, Hu X, Niu H, Tian R, Wang H, Pang H, Jiang L, Qiu B, Chen X, et al. Alterations in the gut microbiome and metabolism with coronary artery disease severity. Microbiome. 2019;7:68. doi: 10.1186/s40168-019-0683-9

8. Schmidt TS, Hayward MR, Coelho LP, Li SS, Costea PI, Voigt AY, Wirbel J, Maistrenko OM, Alves RJ, Bergsten E, et al. Extensive transmission of microbes along the gastrointestinal tract. Elife. 2019;8. doi: 10.7554/eLife.42693

9. Koren O, Spor A, Felin J, Fåk F, Stombaugh J, Tremaroli V, Behre CJ, Knight R, Fagerberg B, Ley RE, et al. Human oral, gut, and plaque microbiota in patients with atherosclerosis. Proceedings of the National Academy of Sciences of the United States of America. 2011;108:4592–4598. doi: 10.1073/pnas.1011383107

10. Jönsson D, Orho-Melander M, Demmer RT, Engström G, Melander O, Klinge B, Nilsson PM. Periodontal disease is associated with carotid plaque area: the Malmö Offspring Dental Study (MODS). Journal of Internal Medicine. 2020;287:301–309. doi: 10.1111/JOIM.12998

11. Ryden L, Buhlin K, Ekstrand E, de Faire U, Gustafsson A, Holmer J, Kjellstrom B, Lindahl B, Norhammar A, Nygren A, et al. Periodontitis Increases the Risk of a First Myocardial Infarction: A Report From the PAROKRANK Study. Circulation. 2016;133:576–583. doi: 10.1161/CIRCULATIONAHA.115.020324

12. Tonetti MS, D’Aiuto F, Nibali L, Donald A, Storry C, Parkar M, Suvan J, Hingorani AD, Vallance P, Deanfield J. Treatment of periodontitis and endothelial function. N Engl J Med. 2007;356:911–920. doi: 10.1056/NEJMoa063186

13. Bergström G, Berglund G, Blomberg A, Brandberg J, Engström G, Engvall J, Eriksson M, Faire Ud, Flinck A, Hansson MG, et al. The Swedish CArdioPulmonary BioImage Study: Objectives and design. Journal of Internal Medicine. 2015;278:645–659. doi: 10.1111/joim.12384

14. Brunkwall L, Jönsson D, Ericson U, Hellstrand S, Kennbäck C, Östling G, Jujic A, Melander O, Engström G, Nilsson J, et al. The Malmö Offspring Study (MOS): design, methods and first results. European Journal of Epidemiology. 2021;36:103–116. doi: 10.1007/s10654-020-00695-4

15. Bergström G, Persson M, Adiels M, Björnson E, Bonander C, Ahlström H, Alfredsson J, Angerås O, Berglund G, Blomberg A, et al. Prevalence of Subclinical Coronary Artery Atherosclerosis in the General Population. Circulation. 2021;144:916–929. doi: 10.1161/CIRCULATIONAHA.121.055340

16. Ohnesorge B, Flohr T, Fischbach R, Kopp AF, Knez A, Schroder S, Schopf UJ, Crispin A, Klotz E, Reiser MF, et al. Reproducibility of coronary calcium quantification in repeat examinations with retrospectively ECG-gated multisection spiral CT. Eur Radiol. 2002;12:1532–1540. doi: 10.1007/s00330-002-1394-2

17. Touboul PJ, Hennerici MG, Meairs S, Adams H, Amarenco P, Bornstein N, Csiba L, Desvarieux M, Ebrahim S, Hernandez RH, et al. Mannheim Carotid Intima-Media Thickness and Plaque Consensus (2004–2006–2011): An Update on Behalf of the Advisory Board of the 3rd and 4th Watching the Risk Symposium 13th and 15th European Stroke Conferences, Mannheim, Germany, 2004, and Brussels, Belgium, 2006. Cerebrovascular diseases (*Basel, Switzerland)*. 2012;34:290. doi: 10.1159/000343145

18. Nielsen HB, Almeida M, Juncker AS, Rasmussen S, Li J, Sunagawa S, Plichta DR, Gautier L, Pedersen AG, Chatelier EL, et al. Identification and assembly of genomes and genetic elements in complex metagenomic samples without using reference genomes. Nature Biotechnology. 2014;32:822–828. doi: 10.1038/nbt.2939

19. O’Leary NA, Wright MW, Brister JR, Ciufo S, Haddad D, McVeigh R, Rajput B, Robbertse B, Smith-White B, Ako-Adjei D, et al. Reference sequence (RefSeq) database at NCBI: current status, taxonomic expansion, and functional annotation. Nucleic acids research. 2016;44:D733–D745. doi: 10.1093/NAR/GKV1189

20. Vieira-Silva S, Falony G, Darzi Y, Lima-Mendez G, Garcia Yunta R, Okuda S, Vandeputte D, Valles-Colomer M, Hildebrand F, Chaffron S, et al. Species–function relationships shape ecological properties of the human gut microbiome. Nature Microbiology. 2016;1:16088. doi: 10.1038/nmicrobiol.2016.88

21. Dekkers KF, Sayols-Baixeras S, Baldanzi G, Nowak C, Hammar U, Nguyen D, Varotsis G, Brunkwall L, Nielsen N, Eklund AC, et al. An online atlas of human plasma metabolite signatures of gut microbiome composition. Nature Communications. 2022;13:5370. doi: 10.1038/s41467-022-33050-0

22. Benjamini Y, Hochberg Y. Controlling the False Discovery Rate: A Practical and Powerful Approach to Multiple Testing. Journal of the Royal Statistical Society Series B (Methodological). 1995;57:289–300. doi: https://doi.org/10.1111/j.2517-6161.1995.tb02031.x

23. Korotkevich G, Sukhov V, Budin N, Shpak B, Artyomov MN, Sergushichev A. Fast gene set enrichment analysis. bioRxiv. 2021:060012. doi: 10.1101/060012

24. Caselli E, Fabbri C, D’Accolti M, Soffritti I, Bassi C, Mazzacane S, Franchi M. Defining the oral microbiome by whole-genome sequencing and resistome analysis: the complexity of the healthy picture. BMC Microbiol. 2020;20:120. doi: 10.1186/s12866-020-01801-y

25. Liu S, Zhao W, Liu X, Cheng L. Metagenomic analysis of the gut microbiome in atherosclerosis patients identify cross-cohort microbial signatures and potential therapeutic target. FASEB Journal. 2020;34:14166–14181. doi: 10.1096/fj.202000622R

26. Doern CD, Burnham C-AD. It’s Not Easy Being Green: the Viridans Group Streptococci, with a Focus on Pediatric Clinical Manifestations. Journal of Clinical Microbiology. 2010;48:3829. doi: 10.1128/JCM.01563-10

27. Lockhart PB, Brennan MT, Sasser HC, Fox PC, Paster BJ, Bahrani-Mougeot FK. Bacteremia associated with toothbrushing and dental extraction. Circulation. 2008;117:3118–3125. doi: 10.1161/CIRCULATIONAHA.107.758524

28. Viehman JA, Smith BJ, Nanjappa S, Li SK, Perez CO, Fernandes CR. Chapter 3 – Microbiology of endocarditis. In: Kilic A, ed. Infective Endocarditis. Academic Press; 2022:43–59.

29. Lanter BB, Davies DG. Propionibacterium acnes recovered from atherosclerotic human carotid arteries undergoes biofilm dispersion and releases lipolytic and proteolytic enzymes in response to norepinephrine challenge In vitro. Infection and Immunity. 2015;83:3960–3971. doi: 10.1128/IAI.00510-15/SUPPL_FILE/ZII999091420SO1.PDF

30. Nagata E, Toledo AD, Oho T. Invasion of human aortic endothelial cells by oral viridans group streptococci and induction of inflammatory cytokine production. Molecular Oral Microbiology. 2011;26:78–88. doi: 10.1111/J.2041-1014.2010.00597.X

31. Hashizume-Takizawa T, Yamaguchi Y, Kobayashi R, Shinozaki-Kuwahara N, Saito M, Kurita-Ochiai T. Oral challenge with *Streptococcus sanguinis* induces aortic inflammation and accelerates atherosclerosis in spontaneously hyperlipidemic mice. Biochemical and Biophysical Research Communications. 2019;520:507–513. doi: 10.1016/j.bbrc.2019.10.057

32. Kesavalu L, Lucas AR, Verma RK, Liu L, Dai E, Sampson E, Progulske-Fox A. Increased atherogenesis during Streptococcus mutans infection in ApoE-null mice. Journal of Dental Research. 2012;91:255–260. doi: 10.1177/0022034511435101

33. Sethi NJ, Safi S, Korang SK, Hróbjartsson A. Antibiotics for secondary prevention of coronary heart disease. Cochrane Database Syst Rev. 2019.

34. Jespersen CM. Randomised placebo controlled multicentre trial to assess short term clarithromycin for patients with stable coronary heart disease: CLARICOR trial. British Medical Journal. 2006;332:22–24. doi: 10.1136/bmj.38666.653600.55

35. Sharma D, Misba L, Khan AU. Antibiotics versus biofilm: an emerging battleground in microbial communities. Antimicrobial Resistance & Infection Control. 2019;8:76. doi: 10.1186/s13756-019-0533-3

36. Xue H, Chen X, Yu C, Deng Y, Zhang Y, Chen S, Chen X, Chen K, Yang Y, Ling W. Gut Microbially Produced Indole-3-Propionic Acid Inhibits Atherosclerosis by Promoting Reverse Cholesterol Transport and Its Deficiency Is Causally Related to Atherosclerotic Cardiovascular Disease. Circ Res. 2022;131:404–420. doi: 10.1161/CIRCRESAHA.122.321253

37. Koh A, Molinaro A, Stahlman M, Khan MT, Schmidt C, Manneras-Holm L, Wu H, Carreras A, Jeong H, Olofsson LE, et al. Microbially Produced Imidazole Propionate Impairs Insulin Signaling through mTORC1. Cell. 2018;175:947–961 e917. doi: 10.1016/j.cell.2018.09.055

38. Bjorne HH, Petersson J, Phillipson M, Weitzberg E, Holm L, Lundberg JO. Nitrite in saliva increases gastric mucosal blood flow and mucus thickness. J Clin Invest. 2004;113:106–114. doi: 10.1172/JCI19019

39. Cosby K, Partovi KS, Crawford JH, Patel RP, Reiter CD, Martyr S, Yang BK, Waclawiw MA, Zalos G, Xu X, et al. Nitrite reduction to nitric oxide by deoxyhemoglobin vasodilates the human circulation. Nat Med. 2003;9:1498–1505. doi: 10.1038/nm954

40. Shiva S, Huang Z, Grubina R, Sun J, Ringwood LA, MacArthur PH, Xu X, Murphy E, Darley-Usmar VM, Gladwin MT. Deoxymyoglobin is a nitrite reductase that generates nitric oxide and regulates mitochondrial respiration. Circ Res. 2007;100:654–661. doi: 10.1161/01.RES.0000260171.52224.6b

41. Christensen SE, Moller E, Bonn SE, Ploner A, Wright A, Sjolander A, Balter O, Lissner L, Balter K. Two new meal-and web-based interactive food frequency questionnaires: validation of energy and macronutrient intake. J Med Internet Res. 2013;15:e109. doi: 10.2196/jmir.2458

42. Silness J, Loe H. Periodontal Disease in Pregnancy. Ii. Correlation between Oral Hygiene and Periodontal Condtion. Acta Odontol Scand. 1964;22:121–135. doi: 10.3109/00016356408993968

43. Langmead B, Salzberg SL. Fast gapped-read alignment with Bowtie 2. Nature Methods 2012 9:4. 2012;9:357-359. doi: 10.1038/nmeth.1923

44. Pasolli E, Asnicar F, Manara S, Zolfo M, Karcher N, Armanini F, Beghini F, Manghi P, Tett A, Ghensi P, et al. Extensive Unexplored Human Microbiome Diversity Revealed by Over 150,000 Genomes from Metagenomes Spanning Age, Geography, and Lifestyle. Cell. 2019;176:649–662.e620. doi: 10.1016/j.cell.2019.01.001

45. Li D, Luo R, Liu CM, Leung CM, Ting HF, Sadakane K, Yamashita H, Lam TW. MEGAHIT v1.0: A fast and scalable metagenome assembler driven by advanced methodologies and community practices. *Methods (San Diego*, Calif). 2016;102:3–11. doi: 10.1016/J.YMETH.2016.02.020

46. Steinegger M, Söding J. Clustering huge protein sequence sets in linear time. Nature communications. 2018;9:2542. doi: 10.1038/S41467-018-04964-5

47. Huerta-Cepas J, Szklarczyk D, Heller D, Hernández-Plaza A, Forslund SK, Cook H, Mende DR, Letunic I, Rattei T, Jensen LJ, et al. eggNOG 5.0: a hierarchical, functionally and phylogenetically annotated orthology resource based on 5090 organisms and 2502 viruses. Nucleic Acids Research. 2019;47:D309–D314. doi: 10.1093/NAR/GKY1085

48. Cantalapiedra CP, Hernández-Plaza A, Letunic I, Bork P, Huerta-Cepas J. eggNOG-mapper v2: Functional Annotation, Orthology Assignments, and Domain Prediction at the Metagenomic Scale. Molecular Biology and Evolution. 2021;38:5825–5829. doi: 10.1093/MOLBEV/MSAB293

