## Supplemental Figures S1-S7 for "*Streptococcus* species abundance in the gut is linked to subclinical coronary atherosclerosis in 8973 participants from the SCAPIS cohort"

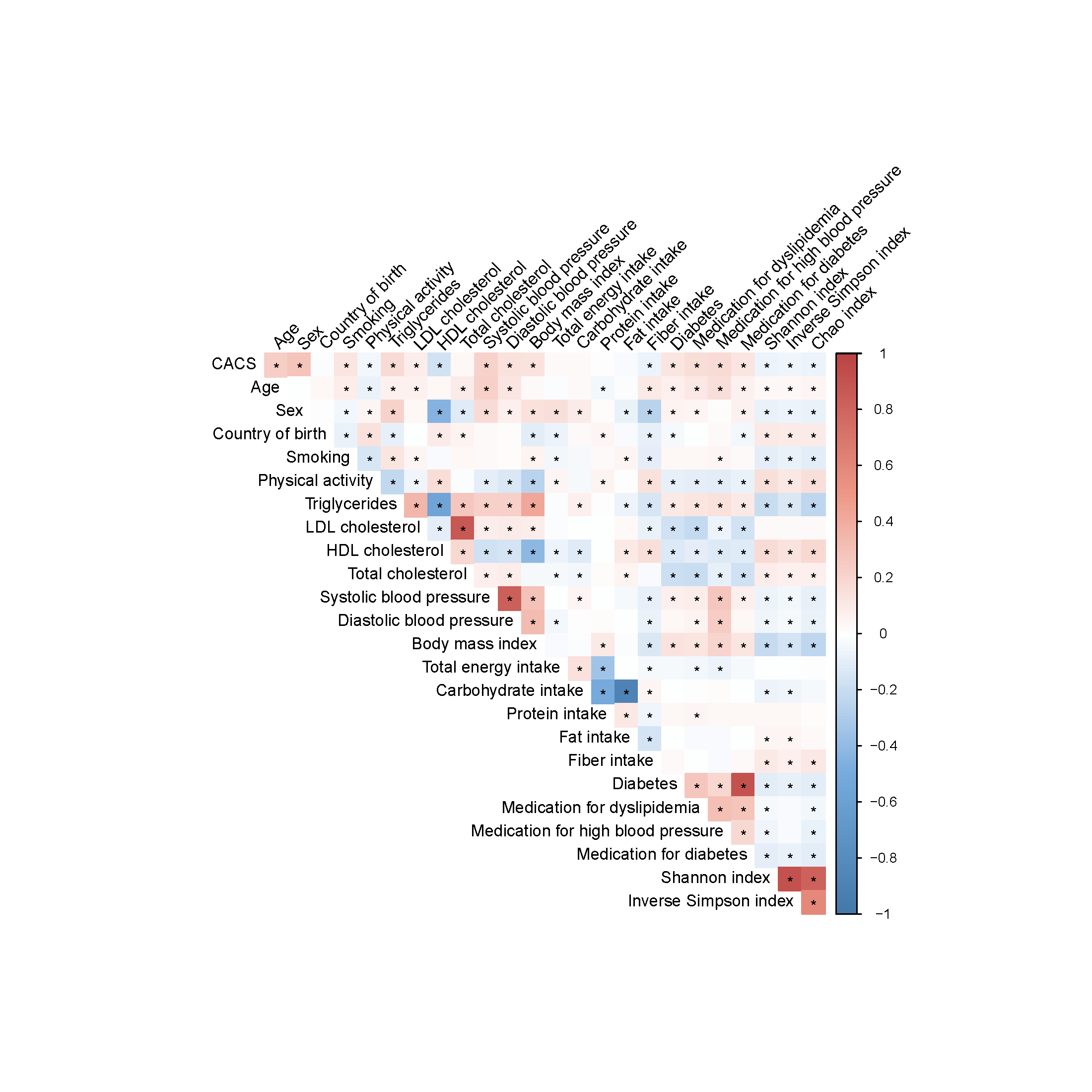

**Figure S1. Spearman correlation of covariates and alpha diversity measurements.** Energy-adjusted intakes of carbohydrate and protein were estimated as percentages of total energy intake. Energy-adjusted fiber intake was estimated as fiber intake per 1000 kcal. **P*<0.05. Country of birth is categorized to Scandinavia/non-Scandinavia.

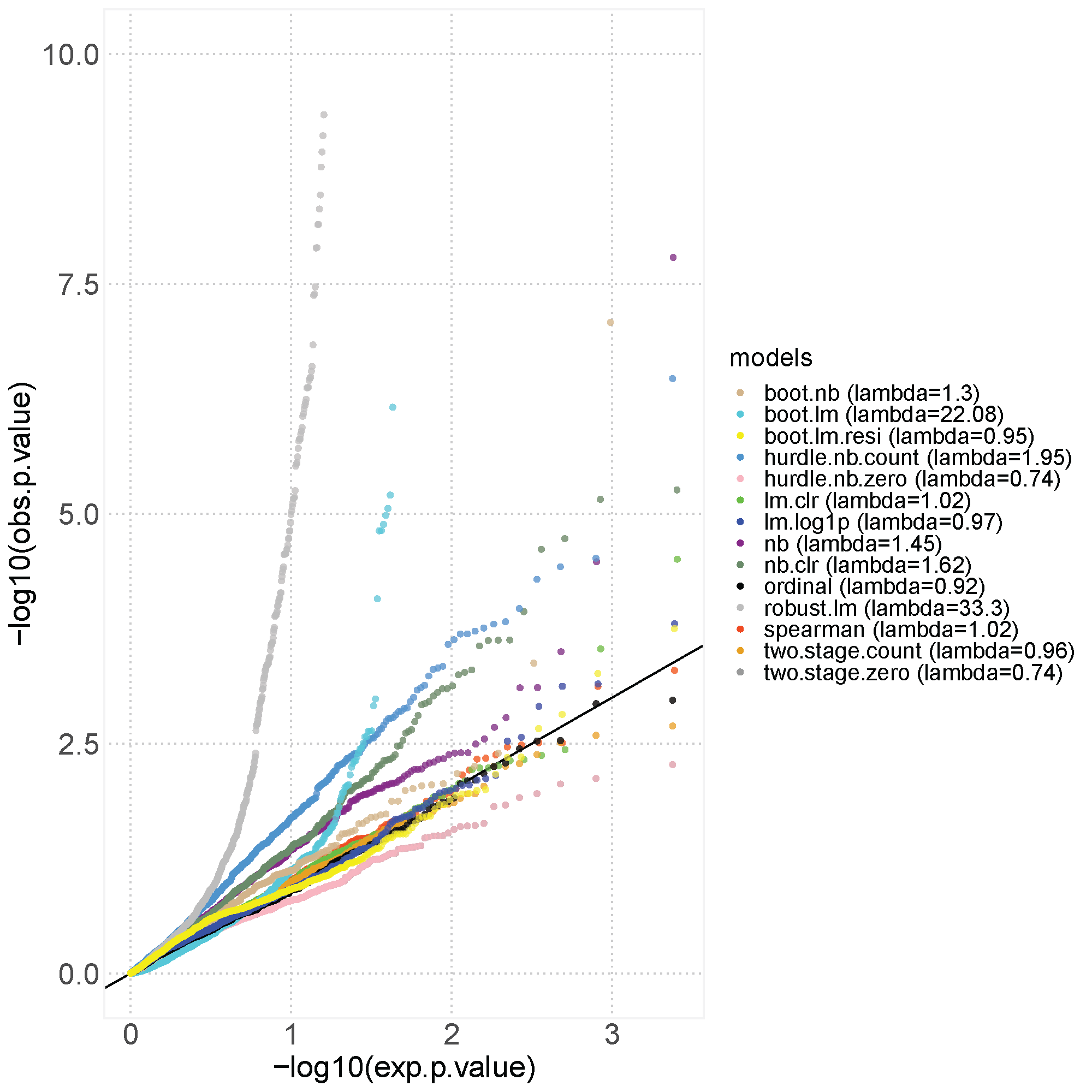

**Figure S2. Quantile-quantile plot (qq-plot) of p-values from simulation studies performed prior to the analysis of the relationship between species and coronary artery calcium score (CACS).** Relative abundances of the first metagenomics data delivery batch (n=438) were shuffled randomly per species to create a simulation dataset with maintained variable distribution. Then, 12 models were tested to compare their performance in the shuffled dataset. Boot.lm, linear model with bootstrapped standard errors; boot.lm.resi, linear model with bootstrapped residuals; boot.nb; negative binomial model with bootstrapped standard errors; hurdle.nb.count, the count part of the hurdle negative binomial model; hurdle.nb.zero, the zero part of the hurdle negative binomial model; lm.clr, linear model with species abundance transformed using the centered log ratio transformation; lm.log1p, linear model with species abundance transformed using the natural logarithm plus one; nb, negative binomial model; nb.clr, negative binomial with species abundance transformed using the centered log ratio transformation; ordinal, ordinal regression model, robust.lm, linear model using robust standard errors; spearman, partial Spearman’s rank correlations; two.stage.count, linear model on the non-zero relative abundances ; two.stage.zero, logistic regression on the species presence or absence. The y-axis is truncated at 10 units to improve the visualization.

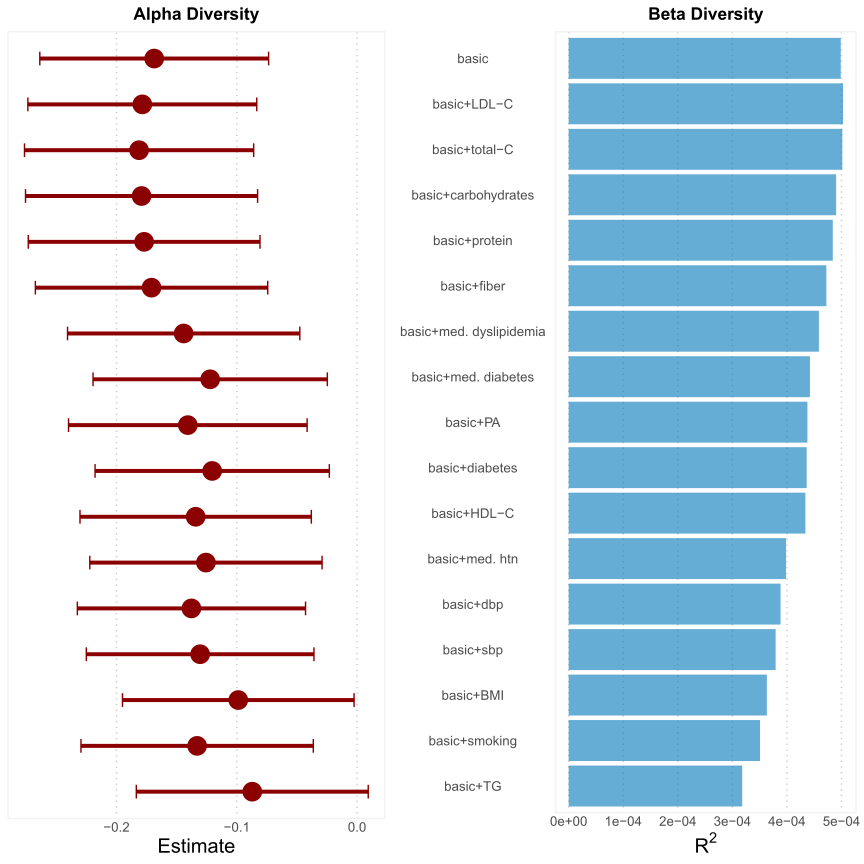

**Figure S3. Addition of each covariate from the full model separately to the basic model to assess the effect on the association between alpha and beta diversity with coronary artery calcium score (CACS).** Alpha diversity was measured using Shannon diversity index and beta diversity using Bray-Curtis dissimilarity. For the analysis of the association between alpha diversity and CACS, the basic model was adjusted for age, sex, country of birth, study center and metagenomics extraction plate as fixed effects, and first-degree family relatedness as random effect (n=8972). For the analysis of the association between beta diversity and CACS, the basic model was not adjusted for family relatedness. Instead, we removed one participant from each family cluster (n=8757). LDL-C: low-density lipoprotein cholesterol; HDL-C: high-density lipoprotein cholesterol, total-C: total cholesterol; TG: triglycerides; sbp: systolic blood pressure; dbp: diastolic blood pressure; PA: physical activity; BMI: body mass index; med. dyslipidemia: self-reported medication for dyslipidemia, med. diabetes: self-reported medication for diabetes; med. htn: self-reported medication for hypertension. The estimations for carbohydrates, protein and fiber were energy-adjusted.

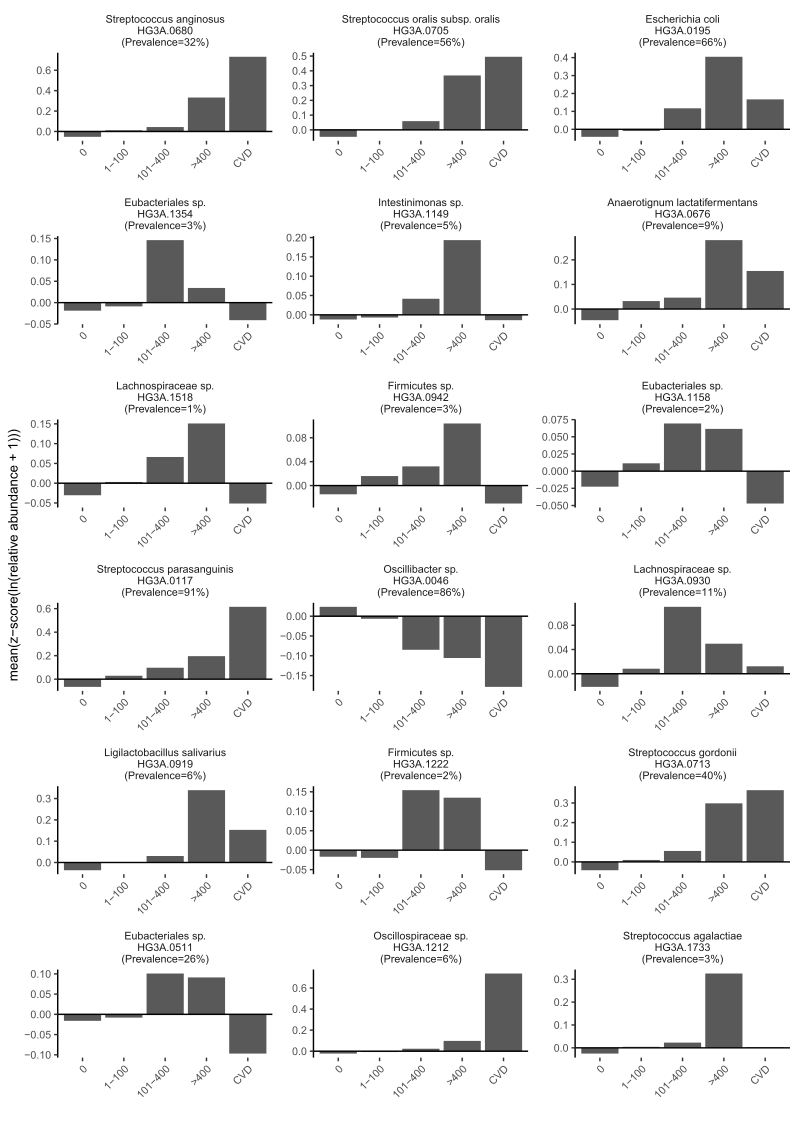

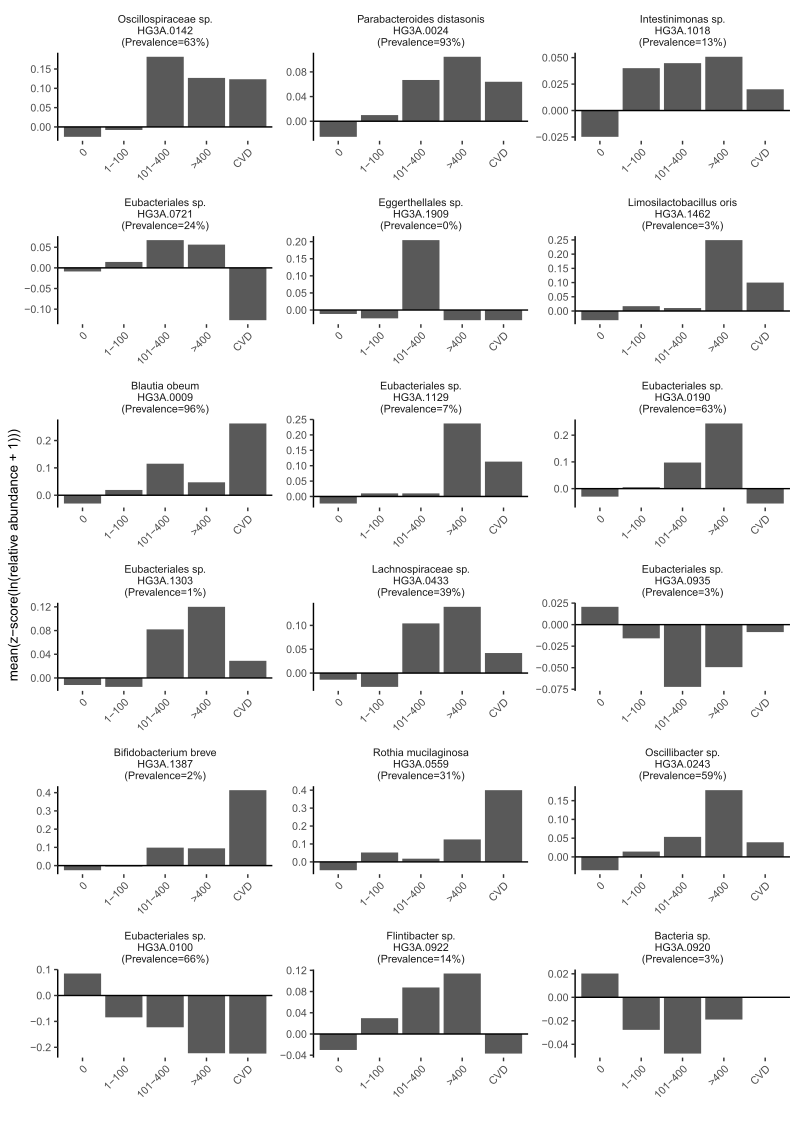

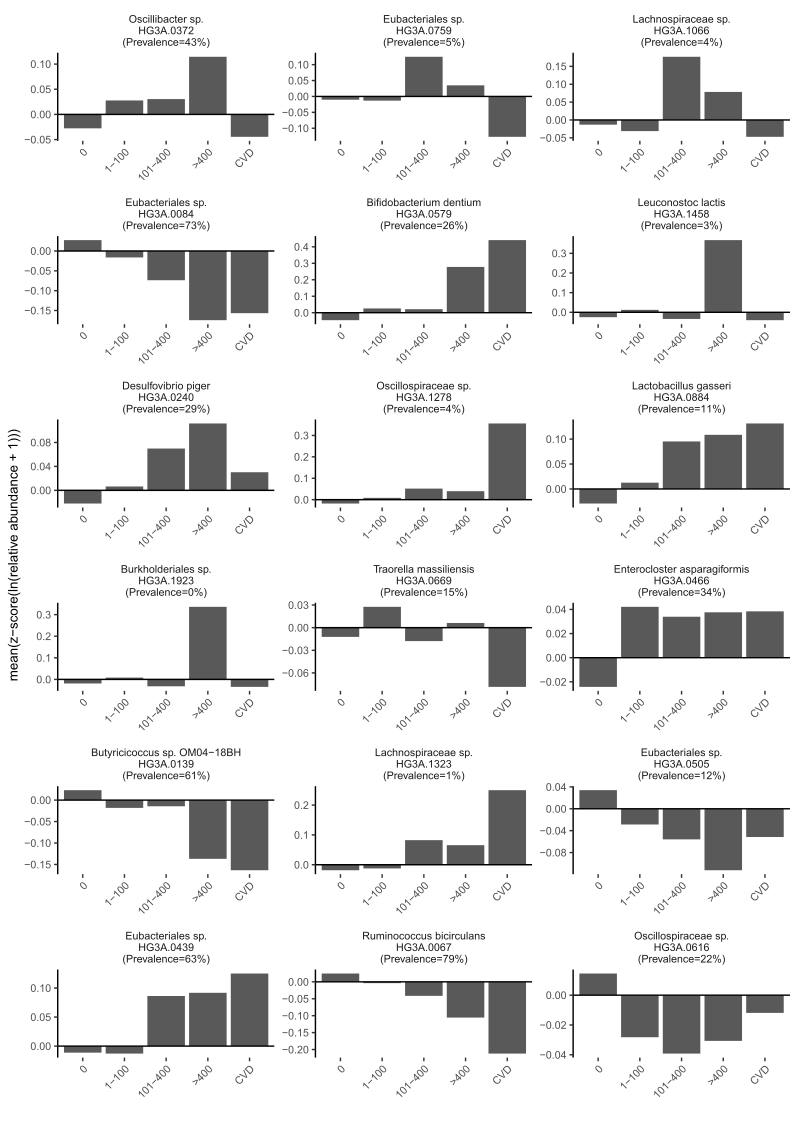

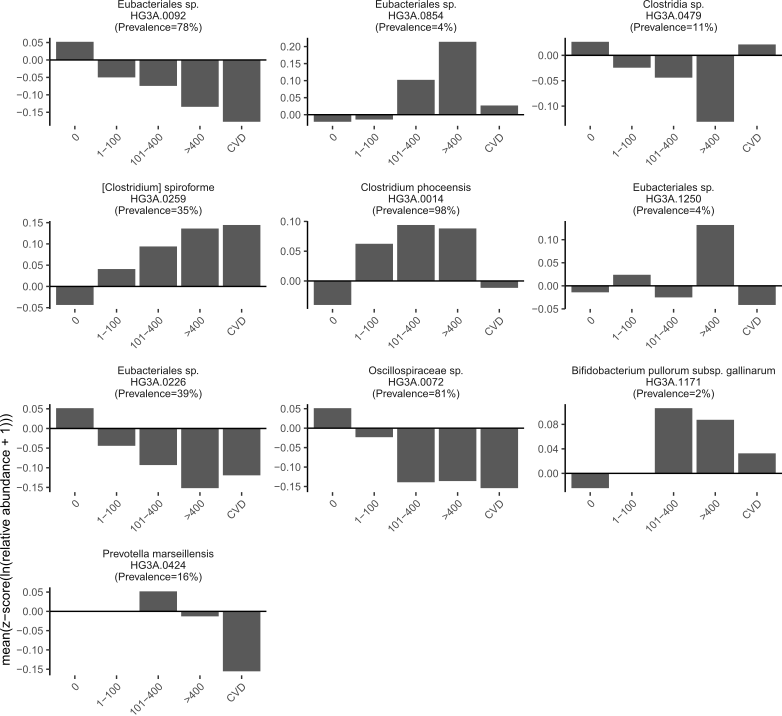

**Figure S4. Relative abundance of 64 CACS-associated species across CACS groups and participants with previous atherosclerotic cardiovascular disease (CVD, n=119), i.e. myocardial infarction, angina, previous bypass surgery or percutaneous coronary intervention, and revascularization of other arterial vessels.**

Species abundance was transformed with the formula ln(x+1), where x denotes the relative abundance of each species. In a second step, a z-transformation was applied to set the mean to 0 and SD to 1.

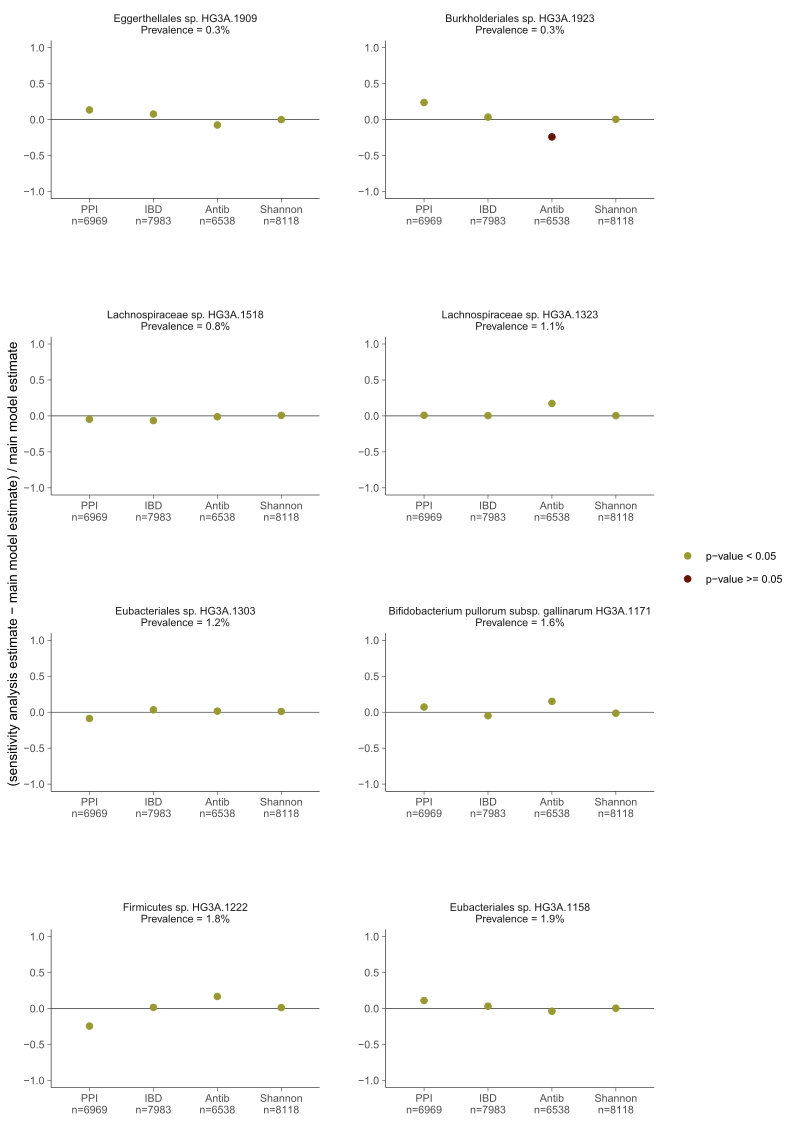

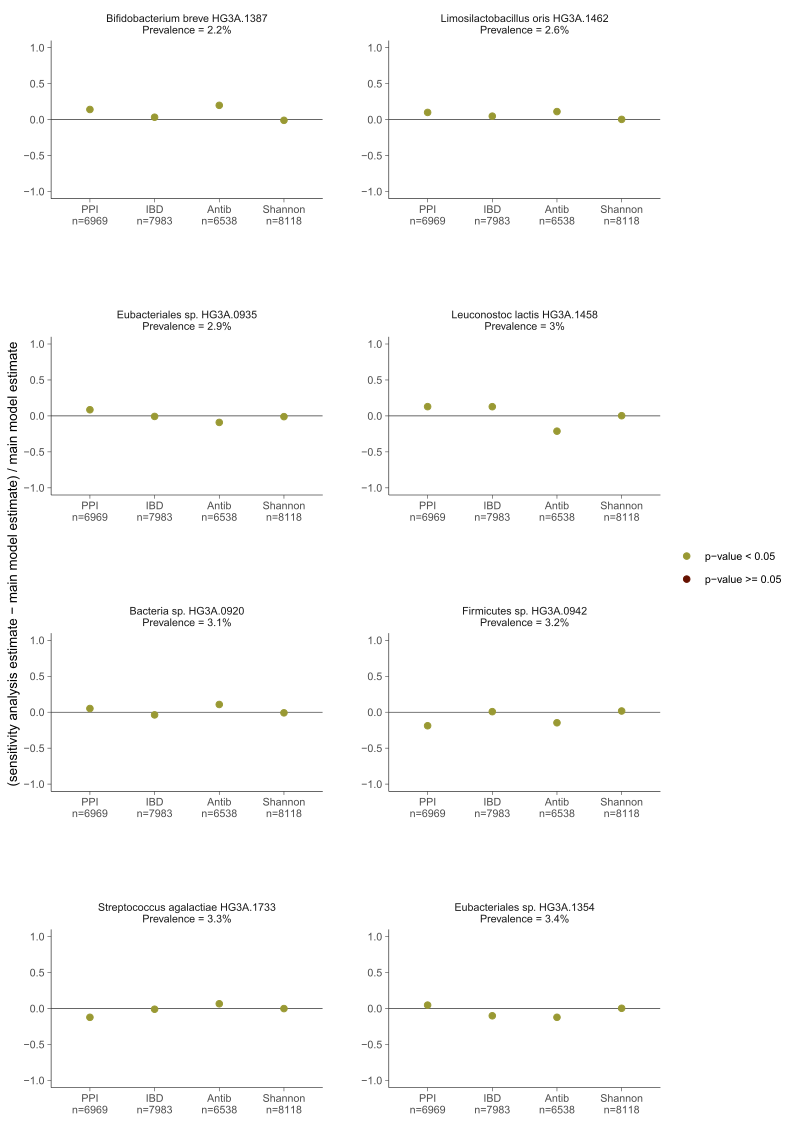

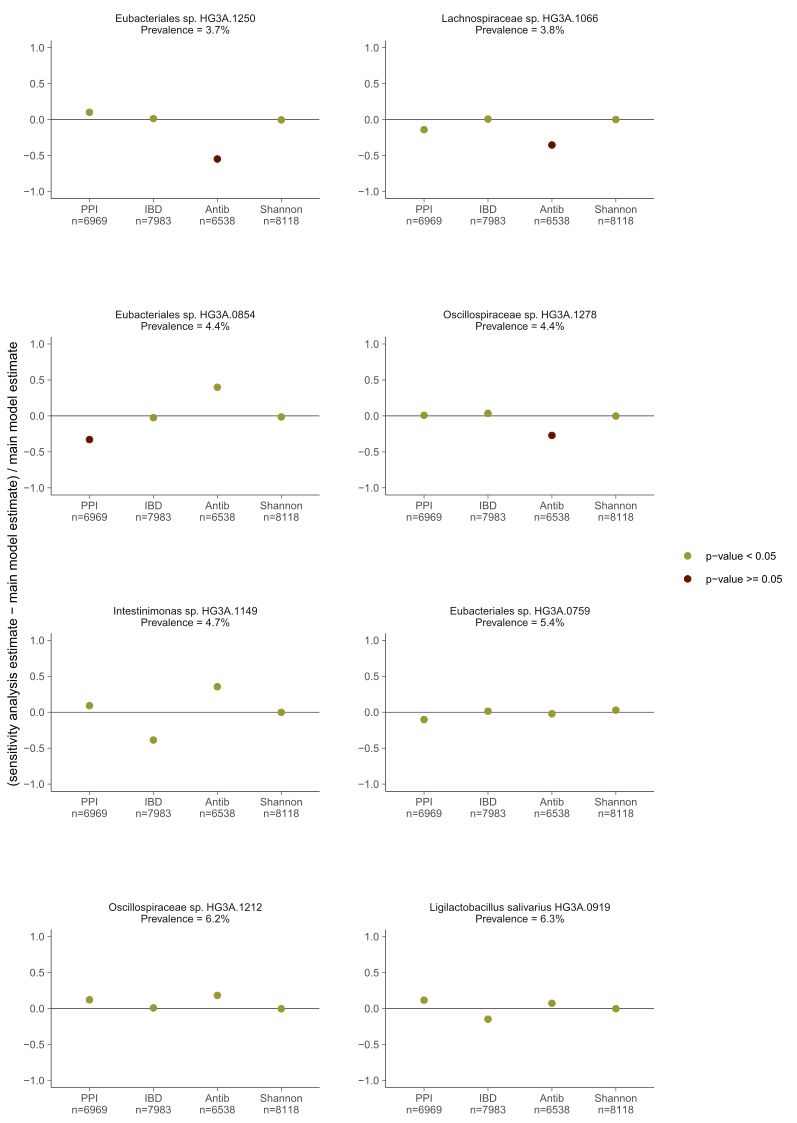

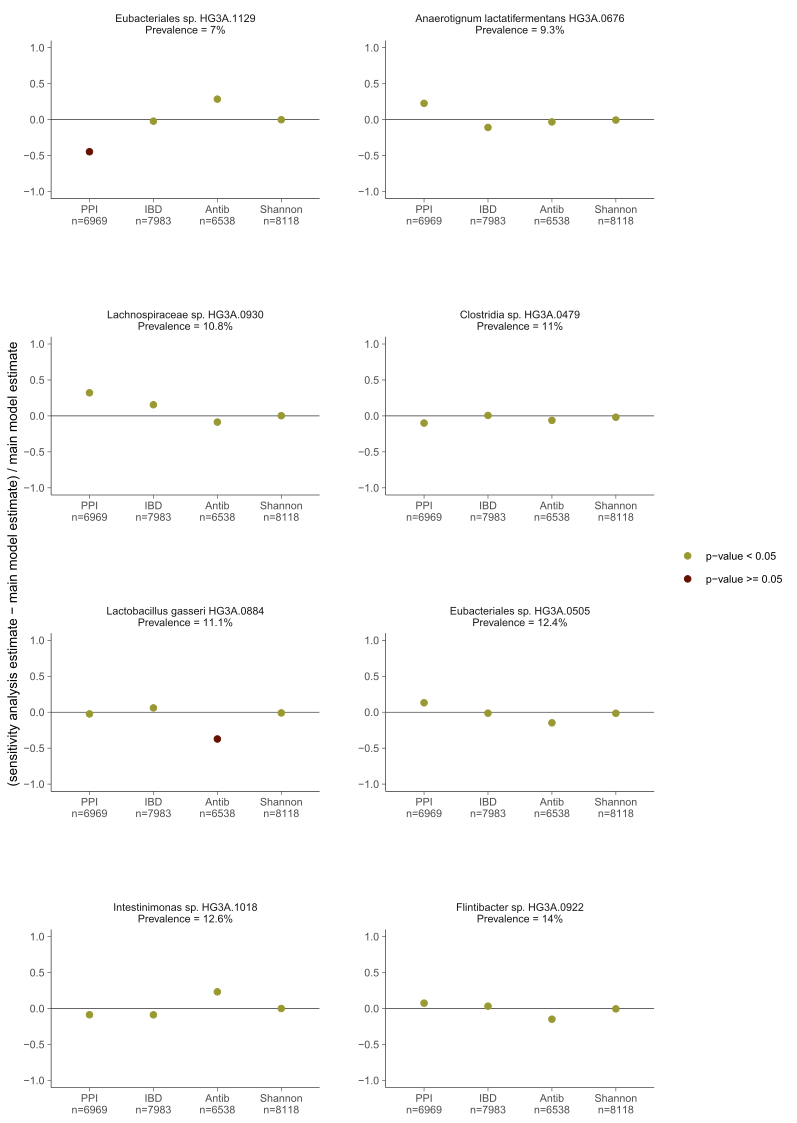

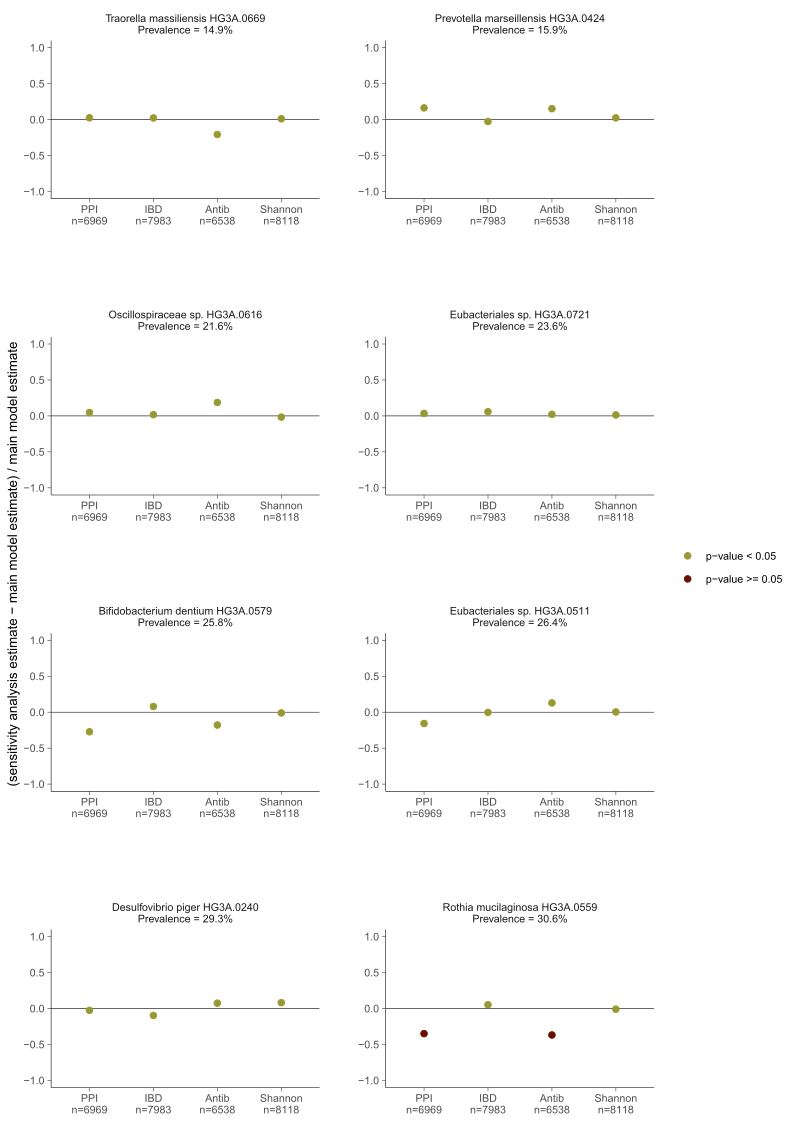

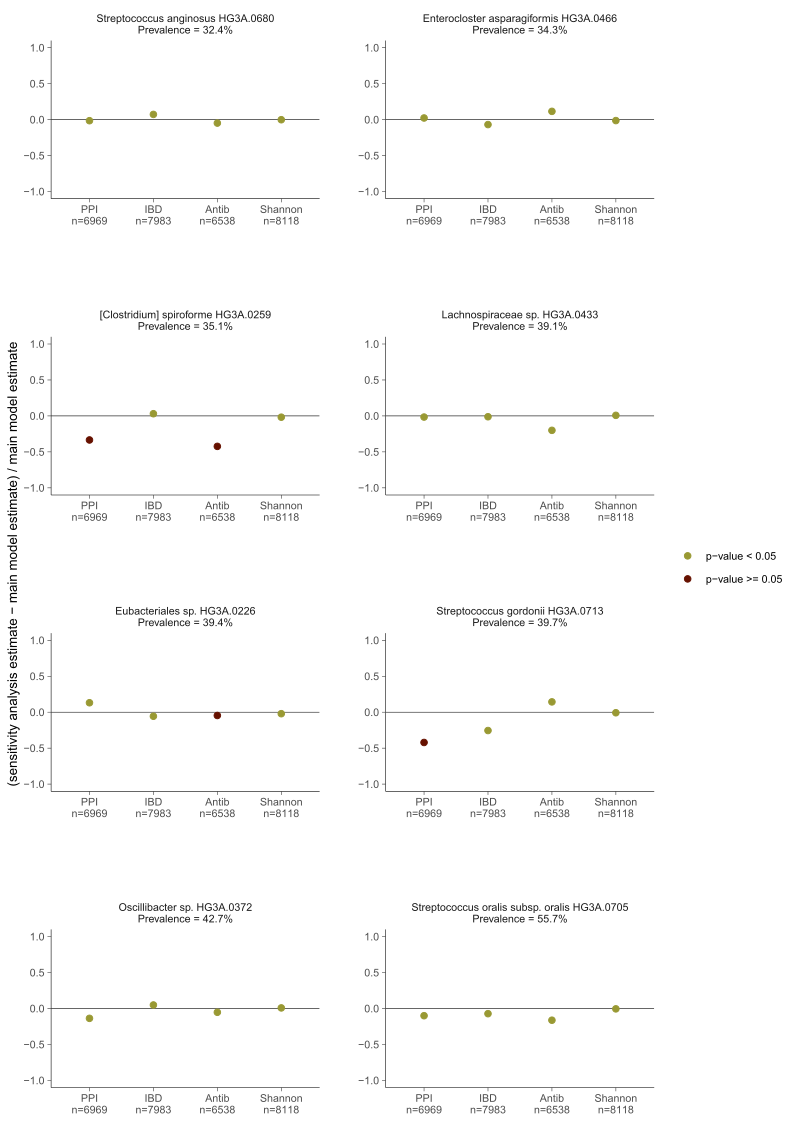

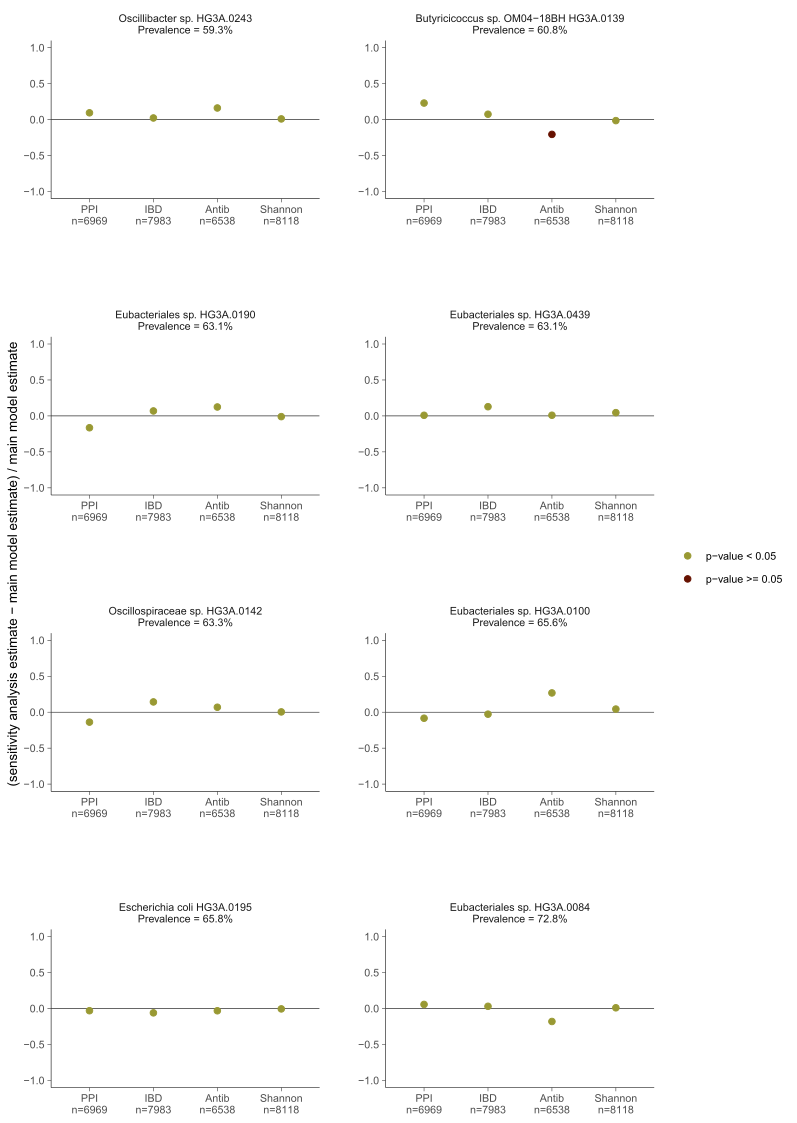

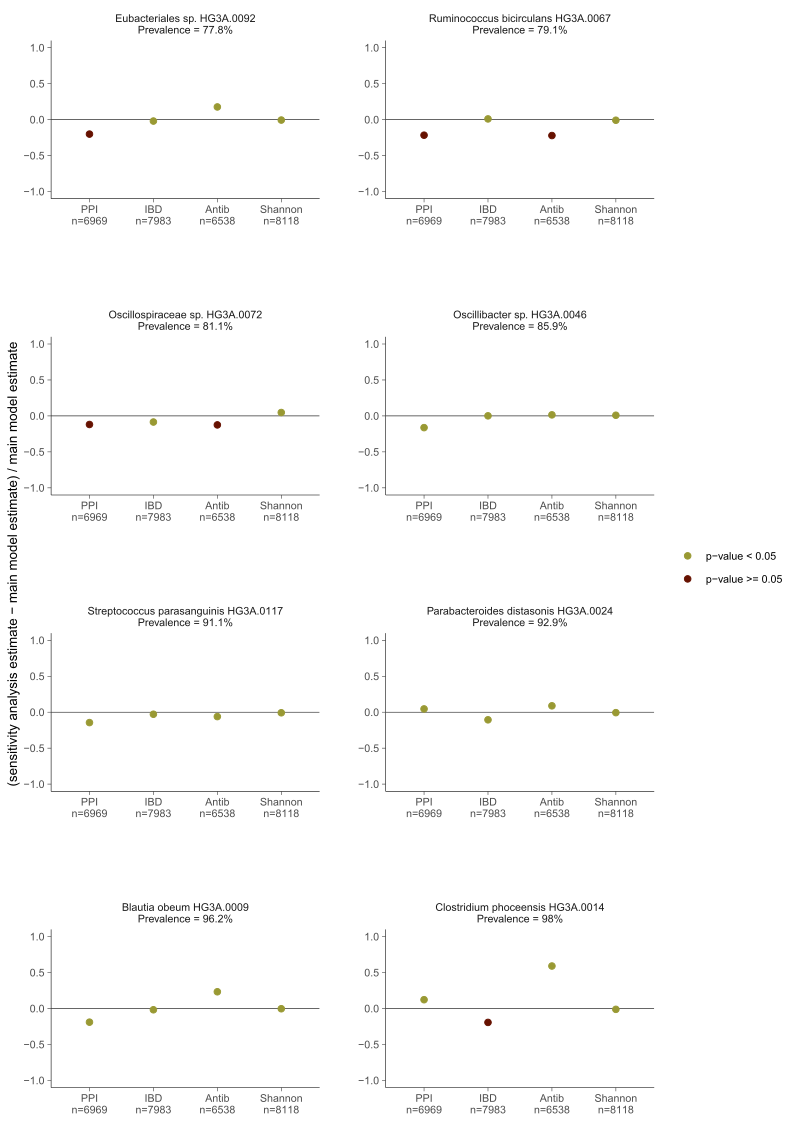

**Figure S5. Sensitivity analysis.**

Association results from the main model in which participants who had proton-pump inhibitors (PPI) measured in plasma, had inflammatory bowel disease, or underwent antibiotic treatment during the year before the baseline visit were excluded, or in which Shannon diversity index was added as a covariate. Main model is adjusted for age, sex, study site, country of birth, metagenomics extraction plate, smoking, physical activity, energy-adjusted carbohydrate, protein and fiber intake, systolic blood pressure, diastolic blood pressure, total cholesterol, high-density lipoprotein, low-density lipoprotein cholesterol, triglycerides, body mass index, diabetes and self-reported medication for dyslipidemia, hypertension and diabetes as fixed effects and first-degree family relatedness as random effect.

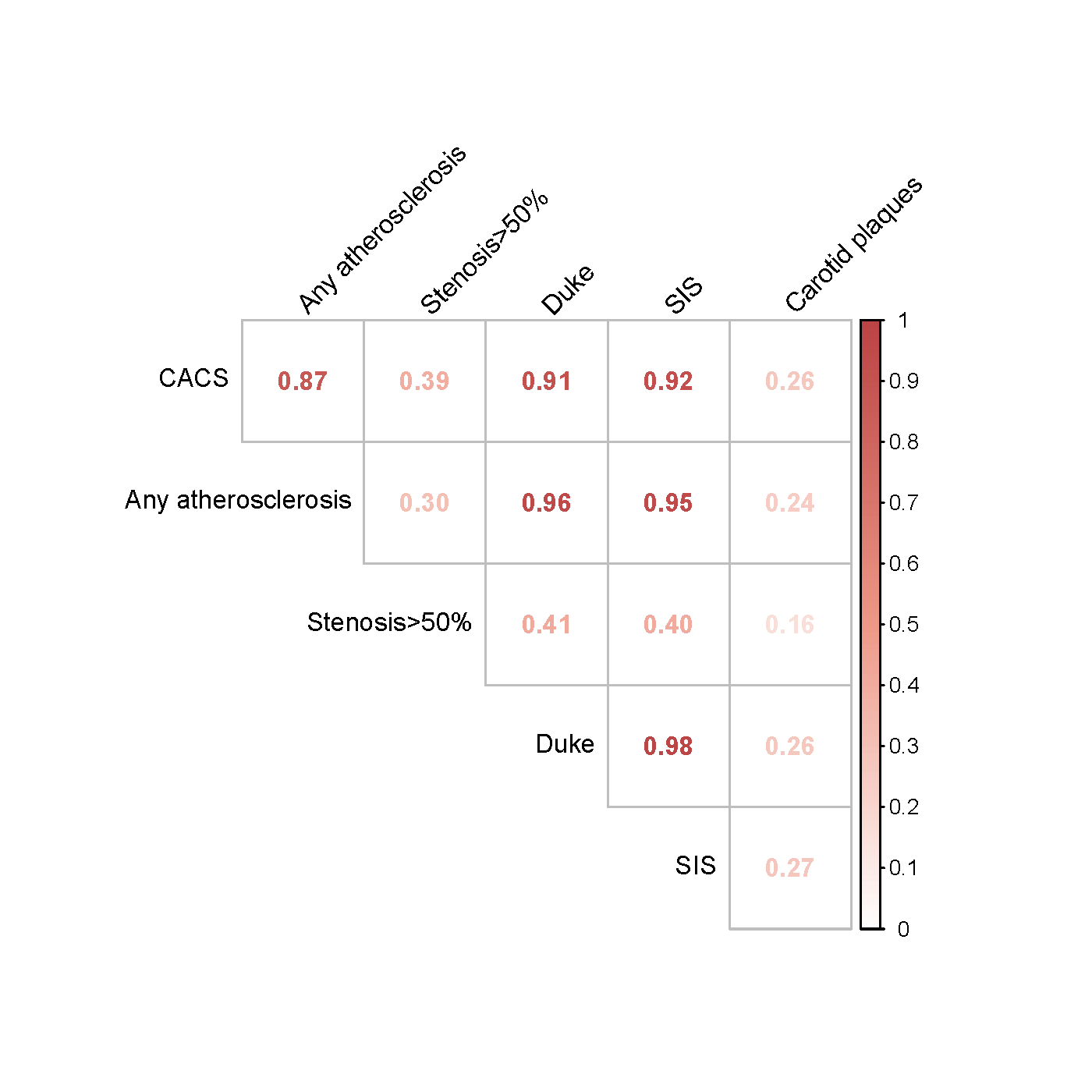

**Figure S6. Correlation of the different atherosclerosis measurements in the SCAPIS study.** CACS: coronary artery calcium score; Any atherosclerosis: any detected coronary atherosclerosis; stenosis>50%: any detected coronary stenosis with an occlusion >50%; Duke: Modified Duke index; SIS: segment involvement score; Carotid plaques: presence of carotid plaques.

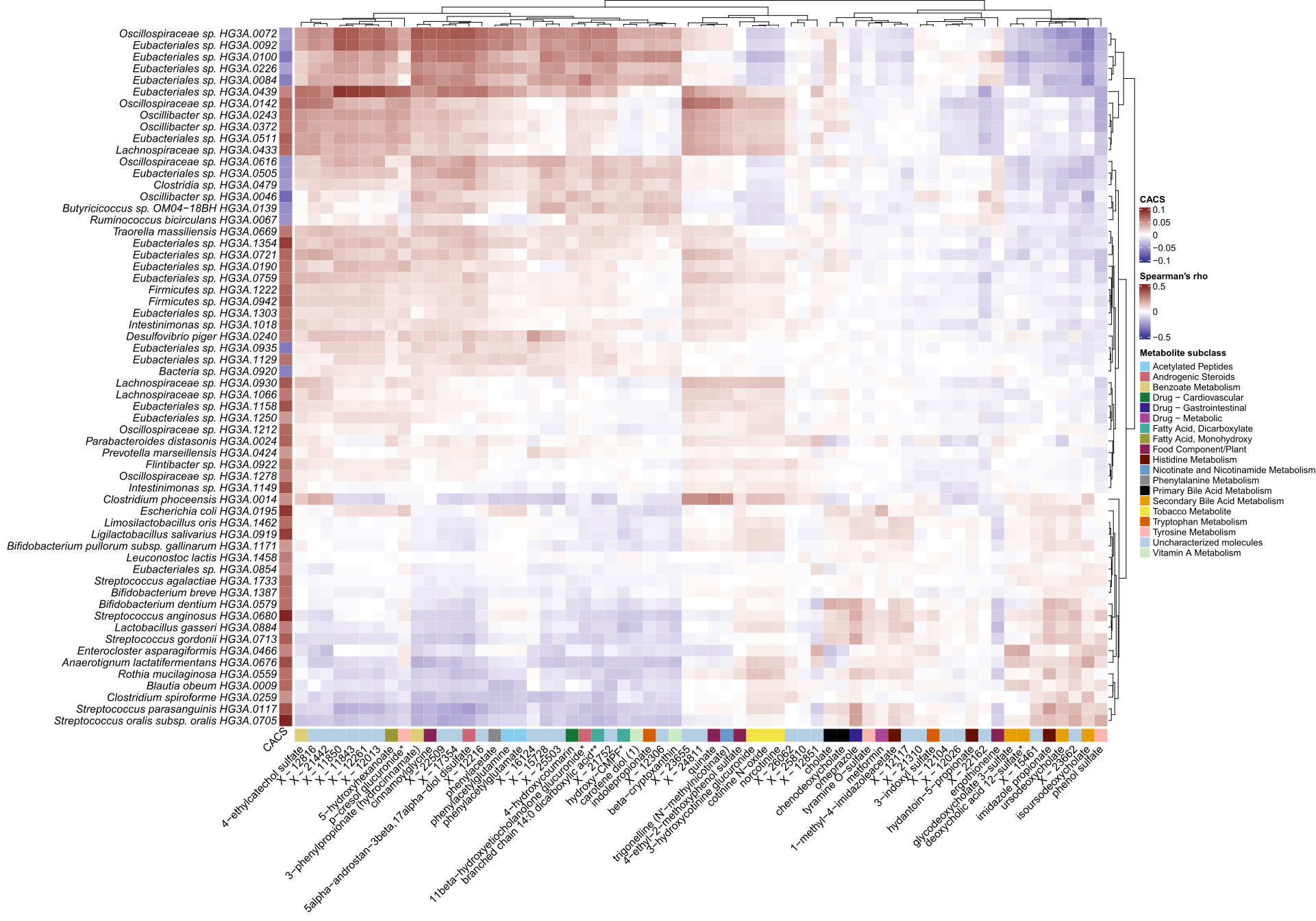

**Figure S7. Heatmap of associations between coronary artery calcium score (CACS)-Associated Species and Plasma Metabolites.** Data were downloaded from the GUTSY Atlas (Supplementary Table 2 and 6, https://gutsyatlas.serve.scilifelab.se/), which is based on the same metagenomics data as the current study. Results were available for 60/64 CACS-associated species. For each CACS-associated species, the three strongest associations with metabolites were selected based on their p-value, and the Spearman’s rank correlation coefficient of the unique subset of 63 metabolites is shown. Hierarchical clustering was performed based on the Euclidian distance. Column CACS is the linear regression coefficient of the species with CACS. The color bar in the bottom depicts the annotated metabolite subclass of the metabolites.
