## Supplemental Methods for "*Streptococcus* species abundance in the gut is linked to subclinical coronary atherosclerosis in 8973 participants from the SCAPIS cohort"

**SCAPIS**

**Study Design and Participants**

The Swedish CArdioPulmonary BioImage Study (SCAPIS) is a national research initiative in Sweden aimed at reducing mortality and morbidity from cardiopulmonary and metabolic diseases. The study was conducted at six university hospitals located in Gothenburg, Linköping, Malmö, Stockholm, Umeå, and Uppsala. ^13^ Individuals 50 to 64 years of age were randomly recruited from the population register, with a participation rate of 50%. In the current study, solely those individuals who provided fecal samples for shotgun metagenomics analysis were considered. These samples were collected from participants recruited at the study sites located in Malmö and Uppsala. Enrollment was initiated in 2014 in SCAPIS-Malmö and in 2015 in SCAPIS-Uppsala in 2015 and completed at both sites in 2018. The study design involved a 3-day-visit scheme for participants, which consisted of a core examination carried out in all the study sites and optional examination that could be added by each site to further their own research interest. The core examination included various assessments, such as anthropometric measurements, lifestyle and dietary questionnaires, blood draws, and imaging using computed tomography (CT), coronary CT angiography, and high-resolution ultrasound, among other procedures. Additionally, optional examinations were available, such as fecal sampling for shotgun metagenomics analysis. Anthropometric measurements, dietary questionnaire, and blood draw were completed at visit 1. Blood pressure was measured at visit 2. Fecal sampling and health and lifestyle questionnaires were completed at home between visits 1 and 2. Once collected, fecal samples were stored at −20°C until visit 2. The median time between visits 1 and 2 was 13 days for Uppsala and 8 days for Malmö. Computed tomography was done at visit 3, a median of 13 days after visit 2. A few fecal samples were returned after visit 2, in Uppsala there were 108 (2%) samples were registered within a week after visit 2 and 39 (0.8%) 8 days or more after visit 2.

**Coronary Atherosclerosis Measurements**

The contrast medium for the CCTA was iohexol (350 mg I/mL; GE Healthcare); the dosage was 325 mg/kg body weight. The images from CCTA were reconstructed and visually scored for atherosclerosis with syngo.via software. The atherosclerosis information from the images was reported using the 18 coronary segment model from the Society of Cardiovascular Computed Tomography. Each coronary segment was visually examined for plaques, and each plaque was characterized. If the CCTA images revealed atherosclerotic plaque in at least one coronary segment, the participant was classified as having coronary atherosclerosis. The segment involvement score (SIS)—the total number of coronary segments with atherosclerotic plaque, was calculated for each participant. The Duke prognostic coronary artery disease index modified for SCAPIS was scored as follows: 0, no atherosclerosis; 1, one stenosis <50%; 2, two stenoses <50% with at least one being proximal or one stenosis >50%; 3, two stenoses ≥50%; 4, three stenoses >50% or two stenoses ≥50% including segment 6; 5, stenosis >50% in at least one of segments 6 or 7 and stenosis ≥50 in at least one of segments 11, 12, or 13 and stenosis ≥50% in at least one of segments 1, 2, 3, or 17; 6, stenosis ≥50% in segment 5. For more information about the CCTA measurements, see Bergström et al. ^15^

**Other Phenotypes**

Age and sex were collected from the Register of the Total Population. Sociodemographic, lifestyle, health, and cardiovascular risk factor information were collected using validated and standardized questionnaires. ^13^ Cardiovascular disease (myocardial infarction, angina, atrial fibrillation, valvular disease, previous bypass surgery or percutaneous coronary intervention, revascularization of other arterial vessel, and stroke), diabetes, and inflammatory bowel disease (Crohn’s diseases and ulcerative colitis) were reported as binary variables by the participant. Extraction plate was categorized in one variable with 112 levels. Self-reported smoking was based on survey question “cqto001 - do you smoke?" with response alternatives (a) never, (b) regular, (c) ex-smoker, (d) occasional, and (e) not willing/able to reply. Regular and occasional smokers combined as current smokers for analysis.

For physical activity in leisure time, participants were categorized as (a) sedentary, (b) moderate exercise, (c) moderate but regular exercise, and (d) regular exercise and training according to self-reported physical activity in question “cqpa12”. Country of birth was categorized as Scandinavia, non-Scandinavian Europe, Asia, and other. The Scandinavia group included participants who were born in Sweden, Denmark, Norway, or Finland. Systolic and diastolic blood pressure were measured in both arms with an automatic device (Omron M10-IT, Omron Healthcare Kyoto, Japan) after 5 minutes of rest in the supine position. The measurements were repeated after a minimum interval of 1 min. The averages of the two systolic and two diastolic blood pressures were calculated in the arm with the highest mean systolic blood pressure.

Total cholesterol, HDL cholesterol, and triglycerides were measured in fresh fasting plasma samples at reference laboratories at each site. LDL cholesterol (was calculated with the Friedewald formula. Triglyceride levels were natural log transformed in the analyses. BMI was determined by dividing the weight (measured in kg) by the square of the height (measured in meters).

In SCAPIS, use of medication for diabetes, hypertension, and dyslipidemia during the preceding 2 weeks was ascertained by questionnaire (yes/no answer). Those participants who had a prescription for antibiotics in the Swedish Prescribed Drug Register during the year preceding the baseline visit were classified as having received antibiotic drug treatment and were further classified as have taken narrow-spectrum and broad-spectrum antibiotics, and could be assigned to both groups. Narrow-spectrum antibiotics included Anatomical Therapeutic Chemical (ATC) codes J01CE02, J01CF05, J01EA01, J01FA01, J01FA06, J01FA09, J01FA10, J01FF01, J01XC01, and J01XE01. Broad-spectrum antibiotics included: J01AA02, J01AA04, J01AA06, J01AA07, J01CA04, J01CA08, J01CR02, J01DB05, J01DD14, J01EE01, J01MA02, J01MA06, J01MA12, J01MA14, J01XX05, and J01XX08. The participants who had measurable omeprazole and/or pantoprazole levels in the plasma over the detection limit were classified as users of proton-pump inhibitors.

Macronutrient intake was assessed with the food frequency questionnaire MiniMeal-Q. ^41^ Energy-adjusted intakes of carbohydrate and protein were estimated as percentages of total energy intake. Energy-adjusted fiber intake was estimated as fiber intake per 1000 kcal. Women who reported values <500 or >5000 kcal/day and men who reported energy intake values <550 or >6000 kcal/day were considered as erroneous reporting and their dietary variables were assigned as missing.

DNA was extracted from whole blood at the Karolinska Institute biobank; genotyping was done at the SNP&SEQ Technology Platform in Uppsala (National Genomics Infrastructure Sweden and Science for Life Laboratory) with a customized version of the Illumina GSA-MDv3. After quality control, kinship among samples was estimated with plink2’s KING implementation; pairs of individuals with KING kinship coefficient >0.177 were assigned as first-degree relatives. The genetic information on 148 samples was not available; those participants were assumed unrelated.

**MOS/MODS**

**Study Design and Participants**

The Malmö Offspring Dental Study (MODS), a substudy of the Malmö Offspring Study (MOS), was carried out in 2013–2021 to identify gene-environment interactions of major diseases. ^10^ The participants were 5259 adults (18 to 71 years of age), which consisted of children and grandchildren from participants examined at the baseline (1992–1996) of the Malmö Diet Cancer Study Cardiovascular Arm. The. The attendance rate of MOS was 47.9%, and details of the study can be found in Brunkwall et al. ^14^ Participants attending MOS in 2014–2018 were eligible to participate in MODS (n=2643) after the second MOS visit. In total 831 individuals were enrolled in MODS.

**Other Phenotypes**

In the MOS population, sociodemographic and lifestyle information and medication treatment were collected with validated and standardized questionnaires. Smoking was categorized as never, former, and current smoker. Education was classified as primary education, secondary education, or university degree.

Information on use of proton-pump inhibitors and antibiotics was obtained from two complementary sources. First, from the National Prescribed Drug Register in Sweden, we acquired information on medications dispensed at a pharmacy during the 12 months immediately before completion of the questionnaire, as judged from ATC codes A02BC (proton-pump inhibitors) and J01 (antibiotics). Second, participants were asked in the questionnaire for ongoing usage (the latest week) of medicines, both prescription and nonprescription, and whether antibiotics had been used the latest 6 months; 88% of the included participants had answered the questionnaire. Participants from MOS were considered as user of a drug if confirmed by the questionnaire or the prescription drug register. For participants in MODS, information was obtained with standardized questionnaires during the dental examination.

**Dental Examination**

In MODS, caries was detected by standard clinical criteria aided by mirror, probe (Hu-Friedy EXD57), and bite-wing radiographs. Cavitated lesions that extended into the dentin were recorded as manifest caries; a primary lesion not reaching the stage of manifest was recorded as initial caries. Initial and manifest lesions were summed for a combined variable (surfaces with caries). Filled surfaces included both fillings and crowns. Both caries and filled surfaces were recorded on all teeth, molars and premolars are considered having five surfaces, front teeth four surfaces. Gingival inflammation was recorded as percentage of bleeding on probing with a Hu-Friedy PCPUNC157 probe, excluding wisdom teeth and counting six surfaces per tooth. Oral hygiene was assessed with the Silness-Löe plaque index but on all teeth excluding wisdom teeth, six sites per tooth. ^42^

**Metagenomics**

*Sample Collection for Shotgun Metagenomics.* In SCAPIS and MOS, fecal samples were self-collected by the participants at home using collection tubes and instruction provided by the SCAPIS center. Participants kept the fecal samples at the home freezer until they attended the study visit. The samples were kept for 7 days at –20ºC in the test centers until they were transported to the central biobank, where they were stored at –80ºC. In MODS, saliva samples were collected by allowing the participant to spit into a 15 mL tube for 5 minutes while chewing a sterile paraffin gum. The samples were immediately placed on ice and quickly stored at –80ºC.

*General Considerations.* For SCAPIS-Malmö and MOS samples, the entire analysis from DNA extraction to relative abundance calculation for each identified species was done with standardized methods at Clinical Microbiomics (Copenhagen, Denmark). SCAPIS-Uppsala samples were analyzed with the same process but the laboratory work was done separately. SCAPIS-Malmö and MOS and SCAPIS-Uppsala, were analyzed separately in random order at the box level. All samples were processed during 2019 and 2020. MODS saliva samples were also analyzed at Clinical Microbiomics following the same pipeline during 2020, but were not processed at the same time as samples from the three other studies. All samples were shipped on dry ice and arrived frozen at the time of delivery to Clinical Microbiomics.

*Handling and Analyses of SCAPIS, MOS, and MODS Samples.* DNA was extracted using NucleoSpin 96 Soil (Macherey-Nagel, Germany). At least one negative control (no sample material) was included per batch of sample during the extraction process. One positive control (Zymogen mock) was included per batch during the whole laboratory process for all the projects, including DNA sequencing. DNA extraction quality was evaluated by agarose gel electrophoresis, and the quantity was determined by Qubit 2.0 fluorometer for the three projects. Genomic DNA was randomly sheared into fragments of approximately 350 base pairs (bp), which were used for library construction with NEBNext Ultra Library Prep Kit for Illumina (New England Biolabs). The sample index pairs were unique for each sample per run. The prepared DNA libraries were purified with the AMPure XP kit and evaluated with an Agilent 2100 Bioanalyzer to determine fragment size distribution. Before sequencing, the concentration of the final libraries was determined by quantitative real-time PCR.

The libraries were sequenced with an Illumina Novaseq 6000 instrument using 2×150 bp paired-end reads. Sequencing generated on average 26.3 million read pairs per sample in SCAPIS-Malmö and MOS, 25.3 million read pairs in SCAPIS-Uppsala, and 26.3 million read pairs in MODS. Reads with >10% ambiguous bases, or >50% bases with Phred score (Qscore) <5 were removed. On average, 97.9% of the sequenced bases had a Qscore >20 in SCAPIS-Malmö and MOS, 97.8% in SCAPIS-Uppsala, and 97.4% in MODS. Reads that mapped to human reference genome GRCh38 using Bowtie 2 v.02.3.4.1^43^ (selecting default settings) were removed from FASTQ files. The remaining reads, classified as high-quality non-host reads (NQNH), were mapped to the gene catalogue using BWA mem v.0.7.16a. The reads were considered mapped if the following criteria were met: an alignment of ≥100 bases, ≥95% identity in this alignment, mapping quality (MAPQ) ≥20, and ≤10 bases failing to align with the gene sequence at either end. Reads meeting previous criteria except the MAPQ threshold were considered multi-mapped. A gene count table was created with the number of mapped read pairs for each gene.

Two specific gene catalogues were built. The first, built for the fecal samples, the human gut catalogue, included 6813 samples from SCAPIS-Malmö and MOS, 4876 from SCAPIS-Uppsala, 9428 from Pasolli et al. ^44^ and 3486 publicly available genome assemblies for isolated microbial strains, selected for their relevance or potential relevance to the human gut or because they are used in commercially available mock microbial communities. The second catalogue, built for the saliva samples, included 706 MODS samples, 1305 oral samples compiled from 21 publicly available data sets, and 1326 publicly available genome assemblies from isolated microbial strains, selected for their relevance or potential relevance to the human mouth, and 81 genome assemblies corresponding to commercially available mock microbial communities.

High-quality non-host reads from the samples were assembled with MEGAHIT (v.1.1.1) ^45^ into contigs of ≥500 bp. The contigs from SCAPIS-Malmö and MOS, SCAPIS-Uppsala, Pasolli et al., ^44^ and genome assemblies were combined, and genes were predicted with Prodigal Gene Prediction Software v.2.6.3 in metagenomics/anonymous mode (https://github.com/hyattpd/Prodigal). The contigs from MODS were combined with genome assemblies, and genes were predicted using the same software. Genes and partial genes with a length <102 bp were removed, resulting in a set of 2.95×10^9^ genes in the human gut catalogue and 1.9×10^8^ genes in the human oral catalogue.

For the human gut catalogue, the gene sequences were split by length into two sets (> 3 kbp and ≤ 3 kbp), and each set was clustered with MMseqs2 (Release 11) (preliminary clustering at 98% identity over 95% coverage of the longer sequence, followed by clustering at 93% identity over 70% coverage of the shorter sequence). For each cluster, a representative sequence was chosen based on the following criteria: first prioritize sequences derived from metagenome assembly (SCAPIS-Malmö and MOS, SCAPIS-Uppsala and Pasolli^44^) over those derived from isolated strain (genomes); then prioritize sequences representing the largest (cardinality) pre-cluster; then prioritize the longest sequence. The two sets of representative sequences were then re-clustered with the same criteria. The resulting sets of short and long cluster representatives were combined as follows. (1) All short cluster representatives were compared to all long cluster representatives and all alignments at 93% identity over 70% coverage of the shorter sequence were identified. (2) All genes that did not have an alignment were retained. (3) For genes that did have an alignment, the short gene but not the long gene was retained. The resulting set of 33.5 million sequences was filtered to retain only sequences that represented a cluster with ≥1 reference-derived sequence and/or ≥5 metagenome-derived sequences, or must have been specifically selected for its relevance, e.g. as a pathogen or as a component of a mock community to build a nonredundant human gut gene catalogue (version “HG3A”) of 14147921 microbial genes.

For the human oral catalogue, the gene sequences were split by length into two sets (> 3 kbp and ≤ 3 kbp), and each set was clustered with MMseqs2 (Release 12) ^46^ at 93% identity over 70% coverage of the shorter sequence. We searched for alignments between long gene cluster representatives and short gene cluster representatives at 93% identity over 70% coverage of the shorter sequence; for all matches, we removed the long gene and retained the short gene. The merged long and short genes were filtered to remove sequences with tetramer entropy below 4, resulting in a nonredundant human oral gene catalogue (version "Ho01") of 8554253 microbial genes.

Metagenomic species (MGS) core gene sets were defined as bins of co-abundant genes identified by gene abundances from the correspondent nonredundant gene catalogue across the cohorts that passed the quality assessment according to Nielsen et al. ^18^ Species abundance was estimated according to the signature gene set, which was assigned using 100 genes with the highest correlation to the median core gene abundance for each species. A table of species counts that considered the total gene counts for the signature gene per species was created. A metagenomics species was considered detected if the read pairs were mapped to least three of the 100 signature genes. Species that did not fulfill this criterion were set to 0, resulting in 99.6% specificity, according to the internal benchmarks. The species count table was normalized for effective gene length (accounting for the read length). The relative abundance of each species was estimated by normalizing it to the sum (100%). All analyses were performed at the species level.

For alpha and beta diversity analyses and descriptive statistics based on median cut-offs, rarefied MGS relative abundance data were used. Rarefied MGS was estimated by random sampling without replacement from the gene count table corresponding to the signature genes. Both SCAPIS-Malmö and MOS and SCAPIS-Uppsala were rarefied to 210430 reads. An outlier sample from SCAPIS-Uppsala was discarded because it had only 1473 reads mapped to the signature genes and rarefying all the samples to 1473 reads would result in a significant loss of sensitivity.

The taxonomical information was annotated after comparison of all the genes on the two catalogues with the NCBI RefSeq database^19^ for archaea, bacterial, fungal, protozoa, and viral genomes, using BLAST algorithms. The human gut catalogue was compared with NCBI RefSeq downloaded on May 2, 2021, and the human oral catalogue was compared with the version downloaded on January 27, 2020. To annotate at the various taxonomic ranks, we required different levels of identity (95%, 95%, 85%, 75%, 65%, 55%, 50% and 45% for subspecies, species, genus, family, order, class, phylum, and superkingdom, respectively) and a minimum of 80% sequence coverage. If >75% of the MGS genes mapped to a single species, the MGS was annotated to this species. For genus, family, order, class, and phylum the thresholds were set to 60%, 50%, 40%, 30%, and 25%, respectively. At the genus and species levels, the MGS was not annotated to this level if >10% of the genes mapped to an alternative species or genus.

The functional annotation was performed by comparing each gene in the catalogue to the EggNOG (v. 5.0) ^47^ orthologous groups database (http://eggnogdb.embl.de/) using EggNOG-mapper software (v. 2.0.1) ^48^ This comparison provided annotation to the Kyoto Encyclopedia of Genes and Genomes (KEGG) orthology (KO) database (https://www.genome.jp/kegg/). The functional potential profile was determined with GMM, ^20^ which includes 103 metabolic pathways that represent a cellular enzymatic process. MGS were assigned to a GMM if they contained at least two-thirds of the KOs required for the functionality of the module. If the module consisted of three or fewer steps, the MGS had to contain all of them. If the module contained alternative paths, the MGS only had to contain one of them.

For SCAPIS-Malmö and MOS, SCAPIS-Uppsala and MODS, no detectable levels of DNA were observed for negative controls, and detectable levels of DNA were observed for mock samples, as expected. The mock samples showed a coefficient of variation, estimated by the Shannon diversity index, of 3.3% in SCAPIS-Malmö and MOS and 3.1% in SCAPIS-Uppsala. The coefficient of variation for 158 pairs of biological replicates randomly introduced in the analysis in SCAPIS-Uppsala center (where Clinical Microbiomics was blind to this information) was 1.5%.

**Statistical Analysis**

*Simulation to determine the main statistical model.* The statistical method for the models with CACS, which was transformed using the natural log plus one transformation except for the negative, ordinal and logistic models, as the outcome was selected based on simulation studies. A simulated dataset was built by randomly shuffling the relative abundances per species for the first batch of delivered data (n=438) to maintain variable distributions and simulate the null hypothesis. Then, 12 models were tested to compare their performance in the shuffled dataset: a linear model with bootstrapped standard errors, a linear model with bootstrapped residuals, a negative binomial model with bootstrapped standard errors, a hurdle negative binomial model, a linear model with species abundance transformed using the centered log ratio transformation, a linear model with species abundance transformed using the natural logarithm plus one, a negative binomial model, a negative binomial with species abundance transformed using the centered log ratio transformation, an ordinal regression model, a linear model using robust standard errors, partial Spearman’s rank correlations, and a two-stage model using linear regression on the non-zero counts and logistic regression on the species presence or absence. The final model was selected according to the performance in the simulation based on the inflation factor, which is a ratio between the median of the empirical distribution of the test statistic and the expected median under the null hypothesis. The linear model with species abundance transformed using a natural logarithm plus one transformation performed well.

*Calculation of partial Spearman correlation from linear mixed models*

For the association of the abundance of gut species with oral species, a partial Spearman correlations from a mixed model based on rank-transformed data was used. Here, we let **y** be a n x 1-sized vector of outcome values, **x** a n x 1-sized vector of exposure values and **C** a n x m-sized matrix of confounder values, where *n* is the number of observations and *m* is the number of confounders. In this case, the partial Spearman correlation between **y** and **x** can be calculated by:

0) rank-transformation of **y**, **x** and **C**
1) regression of **y** on **C**, calculating the residual vector **r_y_**
2) regression **x** on **C**, calculating the residual vector **r_x_**
3) standard correlation analysis between **r_y_** and **r_x_**, obtaining the partial correlation estimate

A p-value with the appropriate degrees of freedom-adjustment can be obtained for this partial correlation by regressing **y** on both **x** and **C** (rank-transformed). This is possible since the p-value for the partial regression coefficient of **x** (**b_x_**) equals the p-value for the partial correlation between **x** and **y**. To generalize the Spearman correlation to a mixed model setting with repeated measures on subject, we modified our procedure:

0) rank-transformation of **y**, **x** and **C**
1) mixed model regressing **y** on **C**, with subject as random effect, calculating the residual vector **r_y_**
2) mixed model regressing **x** on **C,** with subject as random effect, calculating the residual vector **r_x_**
3) standard correlation analysis between **r_y_** and **r_x_**, obtaining the partial correlation estimate.

The p-value for **b_x_** was obtained through a mixed model regressing **y** on both **x** and **C**, with subject as random effect, ensuring that the p-value is adjusted for the proper degrees of freedom as well as the dependence within subjects.
